# Evolution of DNA methylome from precancerous lesions to invasive lung adenocarcinomas

**DOI:** 10.1101/2020.07.11.20142745

**Authors:** Xin Hu, Marcos R. Estecio, Runzhe Chen, Alexandre Reuben, Linghua Wang, Junya Fujimoto, Jian Carrot-Zhang, Nicholas McGranahan, Lisha Ying, Junya Fukuoka, Chi-Wan Chow, Hoa Pham, Myrna C.B. Godoy, Brett W. Carter, Carmen Behrens, Jianhua Zhang, Mara B. Antonoff, Boris Sepesi, Yue Lu, Harvey Pass, Humam Kadara, Paul Scheet, Ara A. Vaporciyan, John V. Heymach, Ignacio I. Wistuba, J. Jack Lee, P. Andrew Futreal, Dan Su, Jean-Pierre J. Issa, Jianjun Zhang

## Abstract

The evolution of DNA methylome and methylation intra-tumor heterogeneity (ITH) during early carcinogenesis of lung adenocarcinoma has not been systematically studied. We perform reduced representation bisulfite sequencing of invasive lung adenocarcinoma and its precursors, atypical adenomatous hyperplasia, adenocarcinoma *in situ* and minimally invasive adenocarcinoma. We observe gradual increase of methylation aberrations and significantly higher level of methylation ITH in later-stage lesions. The phylogenetic patterns inferred from methylation aberrations resemble those based on somatic mutations suggesting parallel methylation and genetic evolution. De-convolution reveal higher ratio of T regulatory cells (Tregs) versus CD8+ T cells in later-stage diseases, implying progressive immunosuppression with neoplastic progression. Furthermore, increased global hypomethylation is associated with higher mutation burden, copy number variation burden and allelic imbalance burden as well as higher Treg/CD8 ratio, highlighting the potential impact of methylation on chromosomal instability, mutagenesis and tumor immune microenvironment during early carcinogenesis of lung adenocarcinomas.

## INTRODUCTION

Lung cancer remains the leading cause of cancer-related death worldwide, yet it is curable if treated early. Many cancers, including lung cancers, are preceded by precancers. Treating precancers to prevent invasive lung cancer is theoretically an attractive approach to reduce lung cancer-associated morbidity and mortality. However, developing strategies for lung cancer prevention has been challenging owing to our limited understanding of neoplastic progression from precancers to invasive lung cancers^1^. Lung adenocarcinoma (ADC) is the most common histologic subtype, accounting for more than 50% of all lung cancers^2^. It has been proposed that invasive lung ADC evolves from atypical adenomatous hyperplasia (AAH), the only recognized precancer for lung ADC, which could progress to ADC *in situ* (AIS), a pre-invasive lung cancer, then to minimally invasive ADC (MIA), and eventually frankly invasive ADC^3^. AAH, AIS, MIA and some ADC often present as pulmonary nodules with a unique radiologic feature termed ground-glass opacity (GGO, hazy nodular opacity with the preservation of underlying bronchial and vascular margins) on CT scans. The biology and clinical course of these lesions are not well defined and their management is controversial. Surgical resection is not the standard of care for treating these lung nodules and the diagnostic yield of biopsy is often low, particularly for GGO-dominant nodules^4^. Therefore, these lung nodules are often referred to as indeterminate pulmonary nodules (IPNs). However, as surgical resection is not often offered, obtaining adequate tissue for comprehensive profiling of these IPNs is difficult, hindering our understanding of the biology underlying these lesions.

We initiated an international collaboration for the collection and characterization of these lung precancers, pre-invasive and early invasive ADC presenting as IPNs. We recently reported the genomic landscape, including the genomic intra-tumor heterogeneity (ITH), and revealed evidence of progressive evolution from AAH to AIS, MIA, and ADC^5^. In addition to mutations, epigenetic alterations such as DNA methylation can also impact neoplastic transformation and fitness. Recent genome-wide methylation profiling studies have revealed that certain alterations, such as the silencing of tumor suppressor genes (TSG) and the activation of genes in stem-like cellular programs^6, 7^ may contribute to carcinogenesis. Our previous study has demonstrated that complex methylation ITH was associated with larger tumor size and increased risk of postsurgical recurrence in patients with invasive lung ADC^8^. Methylation aberrations have been reported in AAH lesions and tumor-adjacent lung tissues suggesting that methylation changes may be early molecular events^9, 10^. However, these pioneer studies only analyzed small numbers of genes implicated in lung carcinogenesis. The dynamic changes in methylome at the genome level and the evolutionary trajectories of methylation ITH during the initiation and progression of lung precancers have not been studied systematically.

In this study, using a unique cohort of resected IPNs of different histologic stages, we delineate the evolution of the methylation landscape and reveal increased methylation ITH in lung ADC than its precursors of early stages, as well as global hypomethylation correlates with immune infiltration, mutational burden and copy number alterations.

## RESULTS

### DNA methylation aberrations increase with the progression of precancers

We performed reduced representation bisulfite sequencing (RRBS) of 62 resected IPNs (14 AAH, 15 AIS, 22 MIA, and 11 invasive ADC) and their paired normal lung tissues from 39 patients (Supplementary Data 1). There was no significant difference regarding age (p= 0.6288, Kruskal-Wallis H test), sex (p= 0.6482, Chi-squared test) or smoking status (p= 0.5696, Chi-squared test) between different histologic groups (Supplementary Data 2). To minimize the impact of “contamination” from non-malignant cells, each IPN specimen was reviewed by two lung cancer pathologists to confirm the diagnosis, mark the areas of diseases and estimate the purity. Only specimens with a minimum of 40% of premalignant or malignant cells were included and manual macro-dissection was applied to enrich for premalignant or malignant cells for DNA extraction. Furthermore, we estimated the tumor purity of these specimens *in silico* based on whole exome sequencing (WES) data by ABSOLUTE^11^. With the caveat that these IPNs had high level of genomic ITH leading to underestimation of tumor purity^5^, we observed no significant difference in tumor purity between different stages (Supplementary Fig. 1).

The mean RRBS sequencing coverage was 58.98 reads. With a minimum of 10 reads shared across all samples, the RRBS profiling allowed the quantification of methylation status at 751,462 CpG sites mapped to 15,761 known genes. Principal component analysis (PCA) demonstrated that the DNA methylome of AAH was more similar to that of normal lung tissue, whereas those of AIS, MIA, and ADC were clearly different from that of normal lung (Fig. 1a). Similarly, unsupervised hierarchical clustering identified two separate clusters: one comprising normal lung and AAH, and the other including AIS, MIA, and ADC (Supplementary Fig. 2). In addition, different regions from the same IPNs tended to cluster together, suggesting that inter-lesion heterogeneity was more prevalent than intra-lesion heterogeneity. Similarly, the methylation profiles of different regions from the same IPNs were highly correlated (median coefficient r=0.884 [range, 0.736-0.969], p < 2.2 x 10^−16^, Supplementary Fig. 3).

**Figure 1.**
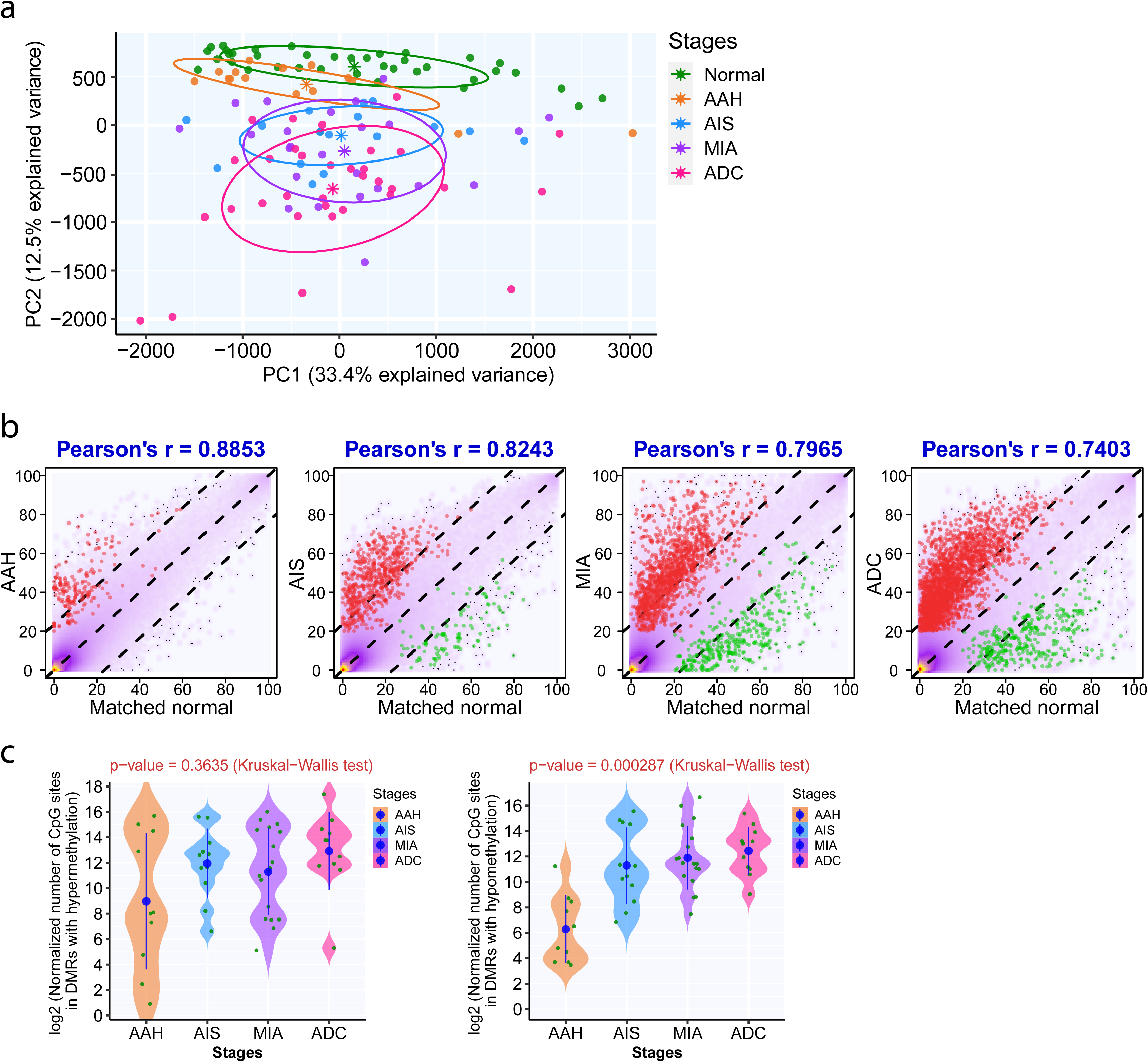
DNA methylation aberrations increase with progression of precancers. (a) Principal component analysis of the methylation profiles of the average methylation values in 39,148 5-kb tiling regions (shared in all samples) composed of autosomal, non-polymorphism CpG sites supported by at least 10X read coverage in IPNs of different stages. The solid dots of different colors represent IPNs of different stages and the star (*) represented the centers of specimens of each histologic stage. (b) Overall genome-wide DNA methylation correlation between IPNs of different stages and matched normal tissue from the same patients. The yellow-purple clouded dots in the smooth scatter plot represent the CpG sites covered by IPN specimens (n = 14 for AAH, n = 11 for AIS, n =18 for MIA, n = 10 for ADC) and paired normal lung. Two-tailed Pearson’s correlation coefficient *r* values are shown on the top. P-values < 2. 2 × 10^−16^ in all 4 histologic stages. The red dots represent CpG sites with hypermethylation in IPNs (within DMRs of methylation gain in IPNs and methylation ≤20% of CpG sites in the corresponding normal lung) and the green dots represent CpG sites with hypomethylation (within DMRs of methylation loss in IPNs and methylation ≥20% of CpG sites in the corresponding normal lung). (c) The number of CpG sites overlapping with DMRs showing hypermethylation (left) or hypomethylation (right) in IPNs of different histologic stages. The green dots represent the mean numbers of CpG sites overlapping with DMRs in each IPN. The solid blue dots represent the mean numbers of CpG sites overlapping with DMRs in IPNs of each histologic stage with 95% confidence interval as error bars. The differences among all stages were assessed using the Kruskal-Wallis H test. (Source data are provided as a source data file.)

There was a progressive increase in the number of CpG sites displaying hypomethylation or hypermethylation (as compared with matched normal lung tissues from the same patients) from AAH to AIS, MIA, and invasive ADC. Hypermethylation appeared to emerge as early as AAH, whereas hypomethylation was evident in only AIS, MIA, and ADC. Meanwhile, the correlation coefficients between the methylation profiles of IPNs and those of their paired normal tissues progressively decreased (r = 0.885 for AAH–normal, 0.824 for AIS–normal, 0.796 for MIA–normal, and 0.740 for ADC–normal, p < 2.2 x 10^−16^, for all comparisons; Fig. 1b). These decreases were marginal reflecting the substantial heterogeneity among different patients and suggesting overall methylation changes may be subtle during early lung carcinogenesis. Nonetheless, these marginal decreases represent methylation alterations in thousands of CpG sites. Further quantification of differentially methylated regions (DMRs) in these IPNs compared with their paired normal lung tissues revealed that later-stage IPNs had numerically more CpG sites with hypermethylation (p = 0.3635, Kruskal-Wallis test) and significantly more CpG sites with hypomethylation (p = 0.000287, Kruskal-Wallis test) (Fig. 1c).

### Evolution of methylome shows similar trend across all genomic regulatory regions

The epigenome is composed of regions with various regulatory elements. To further depict the methylation evolution at different genomic regulatory regions, we first partitioned genomic regions into promoters, enhancers, transcribed regions(transcription) and repressed regions (heterochromatin) based on the peaks of histone marks (H3K04me1, H3K04me2, H3K04me3, H3K09ac, H3K09me3, H3K27ac,H3K27me3, H3K36me3, and H3K79me2) using ChromHMM^12^. We then calculated the numbers of DMRs that overlap with these regions. As shown in Fig. 2a-d, the number of DMRs was higher in later-stage IPNs in all genomic regulatory regions. Next, we selected DMRs overlapping with repetitive elements (UCSC repeatmasker). Later-stage IPNs demonstrated increased DMRs in repetitive regions as well as non-repetitive regions (Fig. 2e, f). Furthermore, as partially methylated domains (PMDs) have been reported to represent a major source of DNA methylation variation in a variety of cancer types^13^, we quantified DMRs inside PMDs versus DMRs outside of PMDs. Similar to the distribution of DMRs in genomic regulatory regions, there were significantly more DMRs in later stage IPNs both inside and outside of PMDs (Fig. 2g, h). Taken together, these results suggested that the majority of methylation alterations during early carcinogenesis of lung ADC may be stochastic across all genomic regulatory regions.

**Figure 2.**
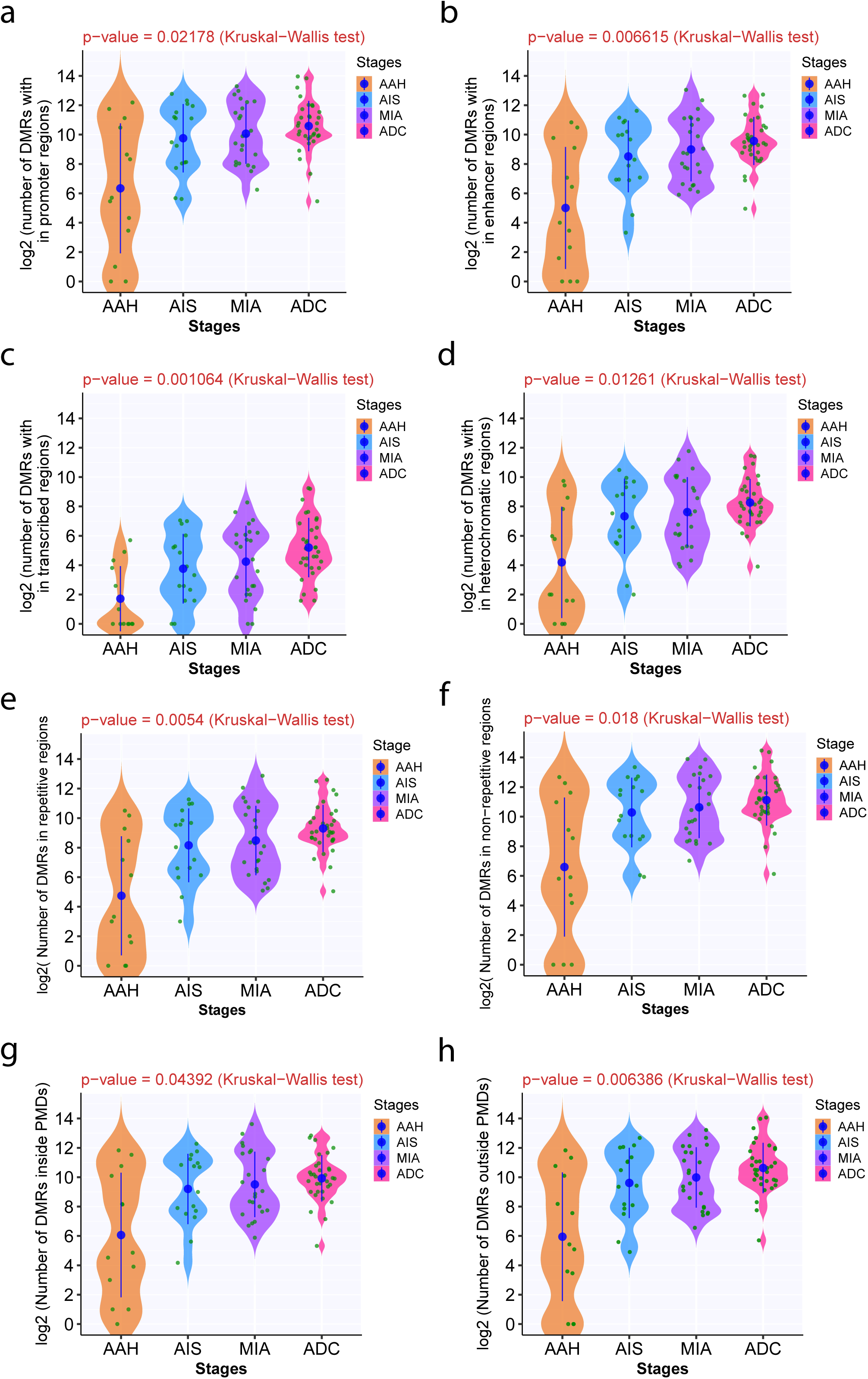
The number of DMRs in IPNs of different stages by genomic regulatory regions and PMDs. The number of DMRs are shown in promoters (a), enhancers (b), transcribed regions (c), repressed (heterochromatic) regions (d), repetitive regions (e), non-repetitive regions(f), inside PMDs (g) and outside PMDs (h). The solid blue dots represent the means of normalized numbers of DMRs in IPNs of each histologic stage with 95% confidence interval as error bars. (Source data are provided as a source data file.)

### Enrichment of transcription factor binding sites at DMRs

DNA methylation can impact the DNA binding of transcription factors (TFs) to their target sequence, termed “motifs”^14^. We searched the motifs that were covered by the DMRs in the IPNs and identified 50 motifs in AAH, 57 in AIS, 64 in MIA, and 66 in ADC that were significantly enriched over all DMRs in each histologic stage (Supplementary Data 3). Many of these motifs were aligned with known binding sites recognized by different TFs known to be involved in critical biological processes, including cell cycle progression, cell proliferation, apoptosis, tumor metastasis, angiogenesis and immune response^15, 16, 17, 18^ (Supplementary Data 4). Motifs associated with *BHE40, BMAL1, CLOCK, EPAS1, MAX, MITF, MXI1, MYC, MYCN, TFE3, USF1* and *USF2* were significantly enriched at DMRs across all stages suggesting the methylation changes at these CpG sites may be early events before the establishment of precancers. On the other hand, some motifs were significantly enriched in DMRs in later-stage IPNs including *ATF3, E2F1, E2F3, E2F4, E2F6, E2F7, EGR1, EGR2, KLF1, KLF12, KLF3, KLF4, KLF6, MYOG, PATZ1, SP1, SP2, SP3, SP4, TFDP1, ZBT14, ZIC1, ZN281, ZN335* exclusively in ADC; *ASCL1, HEN1,KLF9, SNAI2* only in ADC and MIA; *SNAI1* and *TFEB* in ADC, MIA and AIS but not in AAH, suggesting methylation changes at these CpG sites may be associated with later processes during carcinogenesis of these IPNs.

### Later-stage disease has higher methylation ITH

Molecular ITH can have a profound impact on tumor biology^19, 20^, and our previous work demonstrated that methylation ITH was associated with clinical outcome in patients with invasive ADC^8^. To assess the evolution of methylation ITH during the progression of lung precancers, we first calculated the “epiallele shift”, the combinatory difference in epiallele status in all captured loci (>60 reads), for each IPN specimen compared to paired normal lung tissues from the same patients. MIA and invasive ADC showed more pronounced epiallele shifts than AAH or AIS did, indicating a higher level of methylation ITH in IPNs of later histologic stages (Fig. 3a). The corresponding abundance of eloci (loci with distinct epiallele shifts) was also higher in later-stage IPNs (p = 0.004567, Kruskal–Wallis test) (Fig. 3b). When focusing on the 15 patients with multiple IPNs of different stages (IPNs of different stages with the identical genetic background and exposure history), the abundance of eloci was higher in later-stage IPNs than in early-stage IPNs in 13 of the 15 patients (Supplementary Fig. 4), further suggesting that later-stage IPNs have a higher level of methylation ITH. On the other hand, some patients (C23 and J43 for example) had relative high level of methylation ITH in early stage IPNs. Since the post-operative follow up was rather short and the majority of patients are doing well without recurrence, the biological or clinical significance of higher level of methylation ITH in early stage IPNs remains to be determined. Furthermore, we calculated the frequency of 16 combinatory DNA methylation patterns at four consecutive CpG sites covered by the same RRBS reads (termed “epipolymorphism”) in each specimen. Accordingly, later-stage IPNs showed higher epiallele diversity than early-stage IPNs did (Supplementary Fig. 5). Moreover, ADCs had higher levels of epipolymorphism at eloci than MIA, AIS, or AAH did (Fig. 3c), indicating that DNA methylation status had undergone a greater extent of drifting in late-stage IPNs than in early-stage IPNs.

**Figure 3.**
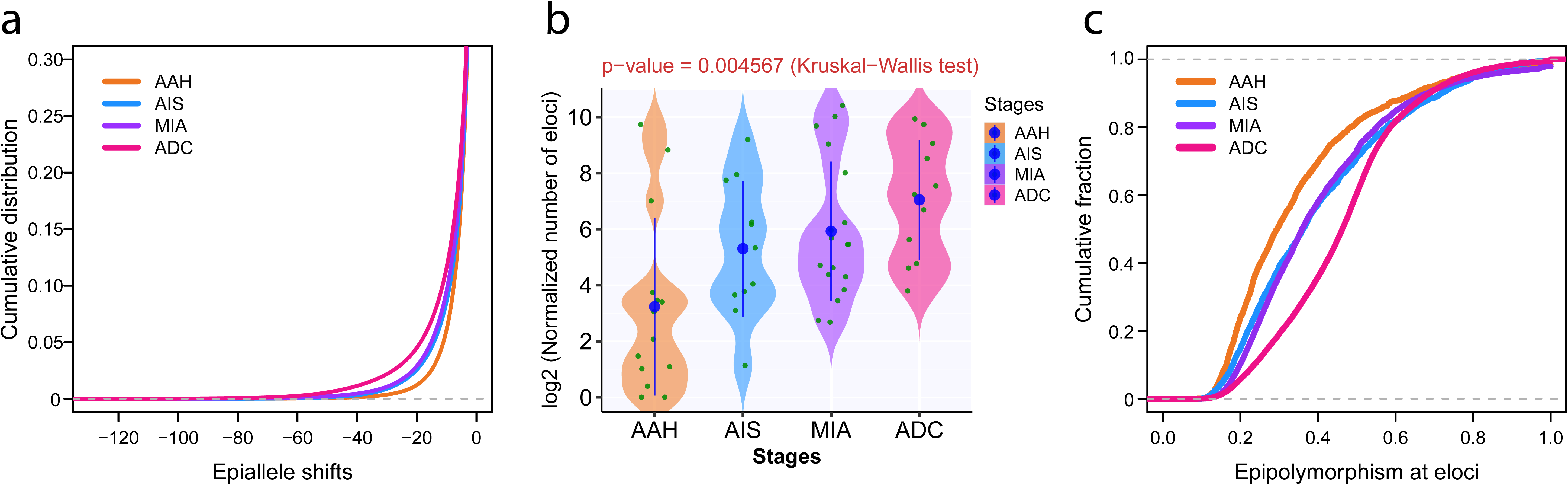
Increased DNA methylation ITH in later-stage IPNs. (a) Cumulative distribution curves of epiallele shifts of the DNA methylome in AAH, AIS, MIA, and ADC specimens compared to normal lung, with 1th percentile of entropy as −21.46 for AAH, −28.88 for AIS, −30.75 for MIA, −42.01 for ADC by quantile regression. (b) The abundance of eloci (loci with distinct epiallele shifts) among IPNs of different histologic stages. Each green dot represents the average number of eloci in each IPN. The solid blue dots represent the mean numbers of eloci in the IPN of each histologic stage with 95% confidence interval as error bars. The differences among all stages were assessed using the Kruskal-Wallis H test. (c) The cumulative distribution curves of epipolymorphism for loci with significant epiallele shifts (*ΔS*< −60) in IPN specimens of AAH, AIS, MIA, and ADC, with 50th percentile of epipolymorphism as 0.295 for AAH, 0.362 for AIS, 0.359 for MIA,0.463 for ADC by quantile regression. *X*-axis denotes the epipolymorphism in IPN specimens at each loci. *Y*-axis denotes cumulative fraction of epipolymorphism from all IPN specimens of each stage. (Source data are provided as a source data file.)

In addition, we sought to assess whether methylation ITH differs at different genomic regulatory regions. As shown in Fig. 4a, loci at heterochromatic regions show the most pronounced epiallele shifts indicating a higher level of methylation ITH at genomic regions of repressed chromatin states. Moreover, there were more eloci inside PMDs than outside of PMDs across all histologic stages (Fig. 4b-e). PMDs are known to be more epigenetically plastic^21^, which makes CpG sites inside PMDs more prone to develop methylation ITH. Additionally, PMDs have been reported to associate with repressive chromatin domains, gene-poor regions and low transcription^21, 22^, therefore, higher level of methylation ITH may be better tolerated inside PMDs.

**Figure 4.**
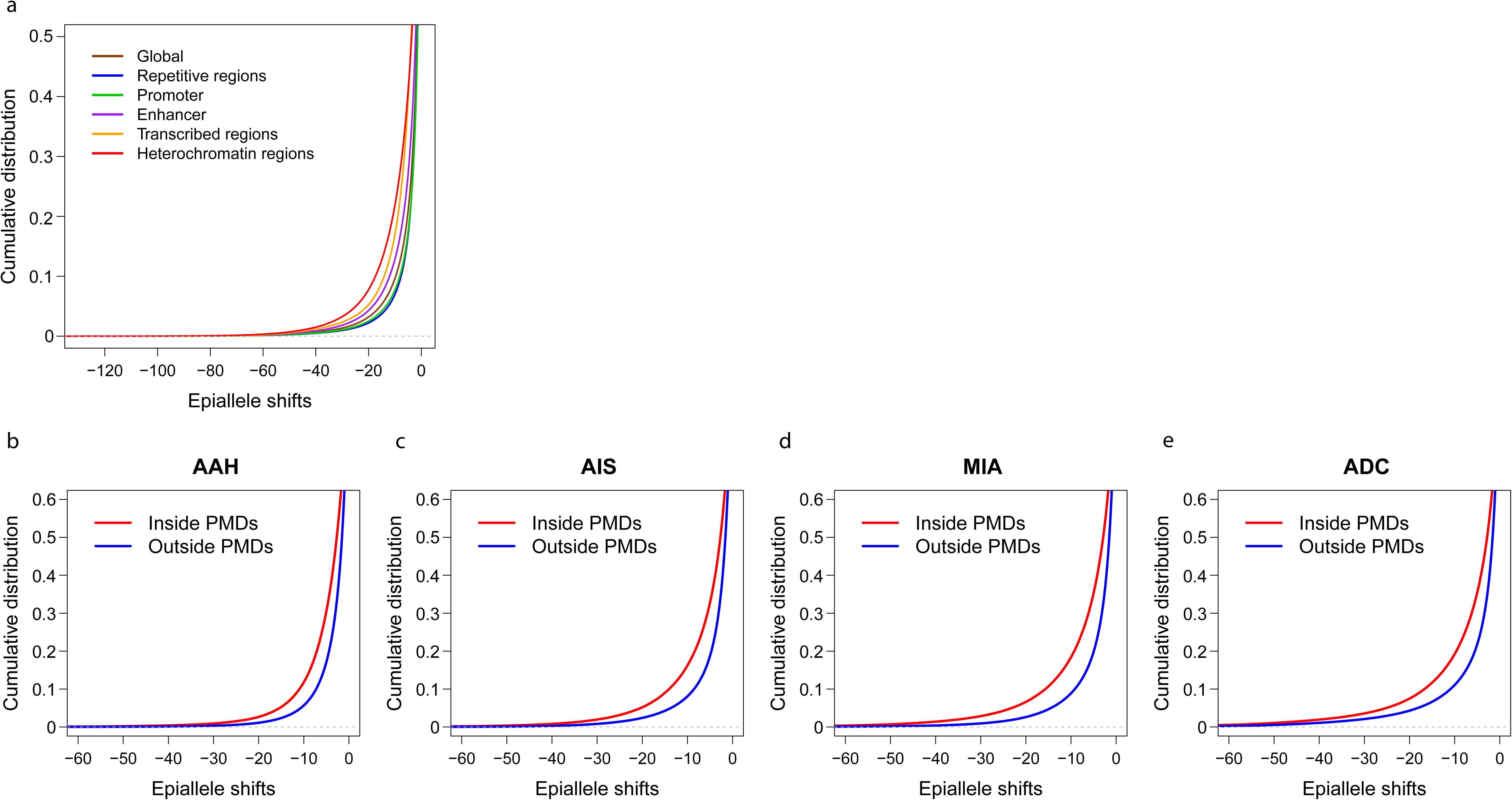
Comparison of methylation ITH by epiallele shifts in different genomic regulatory regions and PMDs. (a) Cumulative distribution curves of epiallele shifts (from tumor sample versus its matched normal sample) of consecutive loci located in promoters, enhancers, transcribed regions, repressed (heterochromatic) regions are shown, with 1th percentile of entropy as −34.07 for all genomic regions, −30.87 for promoters, −38.12 for enhancer, −42.44 for transcribed regions, −45.48 for repressed regions, −29.51 for repetitive regions by quantile regression. Cumulative distribution of epiallele shifts (from IPN sample versus its matched normal sample) of consecutive loci located inside PMDs (red) versus outside PMDs (blue), with 1th percentile of entropy as-29.10 inside PMDs and −20.73 outside PMDs in AAH (b), −37.60 inside PMDs and −28.05 outside PMDs in AIS (c), −44.74 inside PMDs and −29.25 outside PMDs in MIA (d),-50.82 inside PMDs and −41.25 outside PMDs in ADC (e) by quantile regression. The boundary of PMDs are derived from WGBS profiling of normal lung. (Source data are provided as a source data file.)

We next examined the relative distance of eloci to the nearest transcription start sites (TSSs). Interestingly, the vast majority of the eloci were much closer to TSSs in invasive ADC than in AAH, AIS, or MIA (Supplementary Fig. 6). To explore the potential association between methylation ITH and histone modification, we performed Locus Overlap Analysis (LOLA) ^23^ of the genomic regions identified as eloci to evaluate the overlap between genomic regions with methylation ITH and genomic regions targeted by diverse histone posttranslational modifications^24, 25^. LOLA calculates the number of overlapping versus non-overlapping regions to assess the significance of the overlap.After adjustment for false discovery rate considering more eloci in later-stage IPNs, MIA and ADC had higher incidences of eloci significantly enriched in genomic regions occupied by H3K27me3, H3K9me3, and H3K9me2 (Supplementary Data 5), histone modifications strongly associated with transcriptional repression^26, 27, 28^, than AAH or AIS, supporting the concept that histone modifications cooperate with DNA methylation alterations along the evolution of lung precancers^29, 30^.

### Genomic and methylation evolution were primarily in parallel during early lung carcinogenesis

To dissect the evolutionary relationship between the epigenome and genome in lung ADC, we constructed phylogenetic trees. To avoid overfitting, only 5 patients (C5, J7, J8, J9 and J43) with IPNs having a minimum of four spatially separated specimens were included. The overall structure of methylation-based phylogenetic trees was similar to that of phylogenetic trees based on mutations^5^ (Fig. 5a-e). Furthermore, the genomic distance based on somatic mutations was positively correlated with the methylation distance between any pair of specimens from the same IPNs (Fig. 5f) suggesting parallel evolution (either collaboratively or independently) in play during early carcinogenesis of these lung ADCs. Particularly, 4 out of 5 patients with multi-region specimens (C5, J7, J8, J9 and J43) included for phylogenetic analysis demonstrated strong correlation between methylation distance and genomic distance with the only exception of patient J9 (Supplementary Fig. 7). Of note, J9 was a never smoker with the lowest genomic aberration burden (the lowest total mutation burden (TMB), lowest allelic imbalance (AI) burden and the second lowest copy number variation (CNV) burden) and the highest level of genomic ITH (the lowest proportion of trunk mutations) among these 5 tumors, but similar methylation aberration burden comparable to the other 4 tumors (Supplementary Data 6).

**Figure 5.**
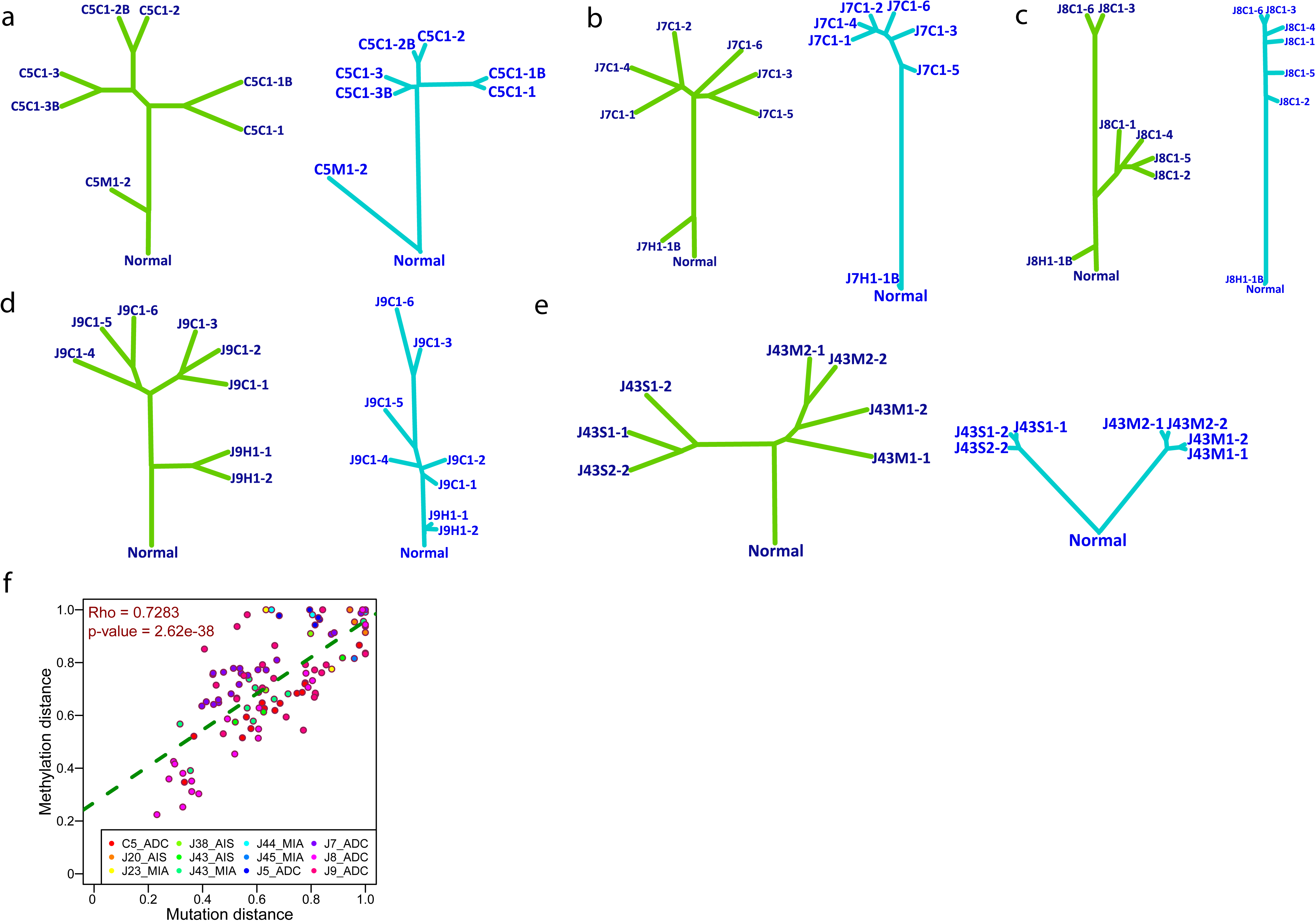
The evolutionary relationship between genomic and methylation landscape. Phylogenetic trees based on mutations (blue) and methylation values (green) in patient C5 (a), J7 (b), J8 (c), J9 (d), J43 (e). The length of each branch indicates the similarity of mutational or methylation profiles between any pair of two spatially separated tumor specimens from each patient. To avoid overfitting, only patients with IPNs having a minimum of four spatially separated specimens were included for this analysis. (f) Correlation of genetic distance (Hamming distance based on all mutations) and methylation distance (Euclidean distance based on methylation values of all CpG sites) between different spatially separated specimens from the same IPNs assessed by two-tailed Spearman’s correlation analysis (p = 2.62 × 10^−38^). Each dot represents the normalized distance between each pair of specimens from the same IPNs. (Source data are provided as a source data file.)

Interestingly, in patient C10, promoter hypermethylation of *TSC2*, a candidate TSG known to inhibit cell growth in lung^31^, was identified in AIS specimens, whereas copy number loss was identified in AAH lesions from the same patient. Similar phenomena of different putative TSGs were observed in several other patients (Supplementary Data 7). Taken together, these data suggested convergent evolution, whereby the same genes or pathways are activated or inactivated by different mechanisms in different cancer cell clones during lung cancer development and progression.

### Global hypomethylation and methylation ITH were associated with increased chromosomal instability in precancerous IPNs

As an essential chemical modification, the methylation status can directly impact the chromosomal structure and DNA mutagenesis. It has been well documented that global hypomethylation is associated with chromosomal instability (CIN) and increased mutational rates in cancers^32, 33^. To further depict the interaction between genome and epigenome during early carcinogenesis of lung ADC, we assessed the global methylation status of these IPNs using long interspersed transposable elements-1 (LINE-1), a widely used surrogate marker for global DNA methylation^34^. As shown in Fig. 6a, we observed a significant decrease of LINE-1 methylation in AIS, MIA and ADC compared to normal lung tissues or AAH indicating increased global hypomethylation in IPNs of later histologic stages. Importantly, methylation level of LINE-1 was inversely correlated with CNV burden (Fig. 6b), AI burden (Fig. 6c) and TMB (Fig. 6d) indicating global hypomethylation is associated with higher level of CIN. Interestingly, LINE-1 methylation status was also inversely correlated with the proportion of clonal mutations in each specimen (Fig. 6e). Furthermore, there was a significant positive correlation between the abundance of eloci and CNV burden as well as AI burden (Supplementary Fig. 8), indicating higher methylation ITH was associated with increased CIN.

**Figure 6.**
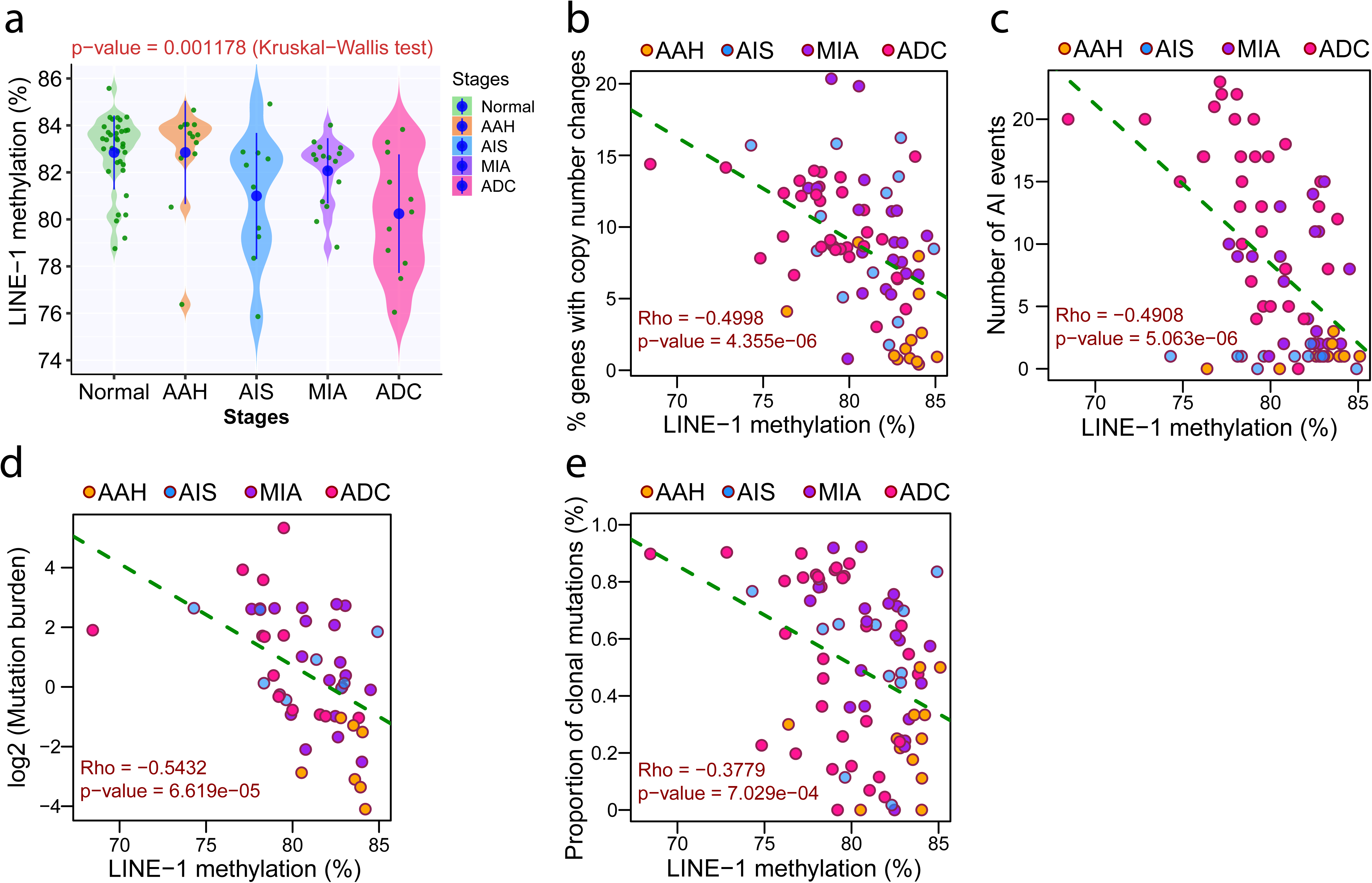
Correlation of LINE-1 methylation with genomic features in IPNs of different stages. (a) LINE-1 methylation level in IPNs of different stages. Each green dot represents LINE-1 methylation level in each IPN and the blue dots represent the means of LINE-1 methylation level in IPNs of each histologic stage with 95% confidence intervals as error bars. The differences among all stages were assessed using the Kruskal-Wallis H test. Correlation between LINE-1 methylation levels and percent of genes with copy number changes (b), number of events with allelic imbalance (AI) (c), mutational burden (log2 transformed) (d), proportion of clonal mutations (e), assessed by two-tailed Spearman’s correlation analysis. Each dot represents each IPN specimen. (Source data are provided as a source data file.)

Given the complex interaction between genome and epigenome, we next categorized CpG sites located in chromosomal regions with various CNV status to assess whether CNV status impacted the results of methylation analyses. As shown in Supplementary Fig. 9, the evolutionary patterns of methylation ITH (represented by the number of eloci) were similar regardless of using CpG sites located in genomic regions with copy number gain, loss or neutral. We also plotted cumulative distribution of epiallele shifts of AAH, AIS, MIA, and ADC using only CpG sites located in copy number-neutral chromosomal regions and observed that later-stage IPNs had higher cumulative distribution of epiallele shifts than early-stage IPNs (Supplementary Fig. 10), similar to that using all CpG sites (Fig. 3a). Furthermore, we regenerated methylation-based phylogenetic trees using only CpG sites located in copy number-neutral chromosomal regions. As shown in Supplementary Fig. 11, the patterns of these methylation-based phylogenetic trees are almost identical to those using all CpG sites. Taken together, these results suggested that in these IPNs, most methylation changes were early molecular events that have occurred before the copy number changes and once established, these patterns were inherited without significant changes from one cell generation to the next.

### Global hypomethylation was associated with suppressed T cell infiltration

Cancer evolution is shaped by interaction between cancer cells and host factors, particularly the host immune response. Given T cells’ central role in anti-tumor immune surveillance^20, 35^, we depicted the T cell infiltration in AIS, MIA and invasive ADC by de-convoluting RNAseq data from an independent dataset recently published^36^. Our analysis demonstrated an increase of CD4+ T regulatory cells (Tregs) (p=4.25e-27) and decrease of CD8+ T cells (although the difference was not significant, p=0.1374) from normal lung tissues to AIS/MIA and invasive ADC leading to significantly higher Treg/CD8 ratio in invasive ADC (p=3.17e-31) (Supplementary Fig. 12a-c). We next applied MethylCIBERSORT^37^ to delineate the T cell infiltration of IPNs in our cohort. Similarly, we observed higher infiltration of Tregs (Fig. 7a, p=0.009758) and lower infiltration of CD8+ T cells in later-stage IPNs although the difference did not reach statistical significance (Fig. 7b, p=0.1472), leading to significantly higher Treg/CD8 ratio in later-stage IPNs (Fig. 7c, p=0.00194). As increased Treg/CD8 ratio is known to associate with suppressed anti-tumor immune surveillance^38^, these results indicated more suppressive immune microenvironment in later stage IPNs, consistent with our previous findings^39^. Interestingly, Treg/CD8 ratio was inversely correlated with LINE-1 methylation level (Fig. 7d) implying the potential association between global hypomethylation and immunosuppression.

**Figure 7.**
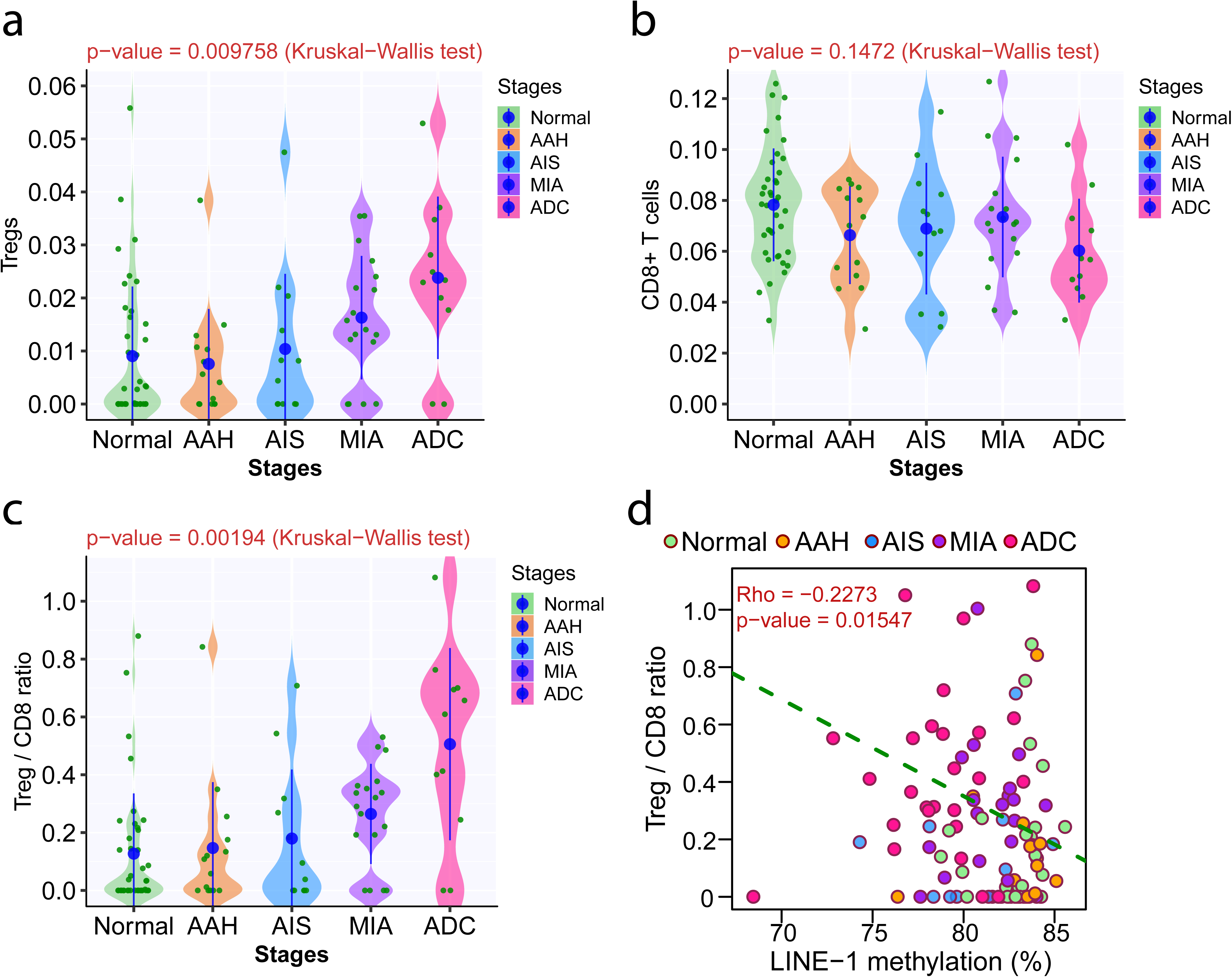
T cell infiltration in IPNs of different stages. The immune cell fraction of T regulatory cells (Tregs) (a), CD8+ T-cells (b), Treg/CD8 ratio (c) in IPNs of different stages. Each green dot represents immune cell fraction in each IPN and the blue dots represent the means of immune cell fraction in IPNs of each histologic stage with 95% confidence intervals as error bars. The differences among all stages were assessed using the Kruskal-Wallis H test. Infiltration of T lymphocytes was inferred by MethylCIBERSORT. (d) Correlation between LINE-1 methylation and Treg/CD8 ratio was assessed by Spearman correlation analysis. Each dot represents each specimen. (Source data are provided as a source data file.)

## DISCUSSION

The methylation landscape has been studied extensively in various malignancies^40, 41, 42^. However, methylation aberrations in precancers are poorly defined, largely due to lack of appropriate specimens. Using small panels of genes implicated in lung carcinogenesis, previous studies have demonstrated gradual changes in DNA methylation in AAH, AIS, and ADC^9, 10^. However, due to small number of loci, primitive technology and lack of genomic data, many critical questions regarding methylation evolution during early carcinogenesis were not addressed. Leveraging a unique collection of resected IPNs of different stages, we delineated the DNA methylome in lung precancers, preinvasive ADC and invasive ADC. Our results demonstrated an increase in both hypermethylation and hypomethylation in later-stage diseases. Interestingly, compared to normal lung tissues, hypermethylation appeared to emerge as early as at AAH, whereas hypomethylation only became obvious after AIS (Fig. 1b), implying that somatic hypermethylation may have preceded hypomethylation during early carcinogenesis of lung ADC.

Different cells within the same tumor can exhibit different molecular and phenotypic features, a phenomenon termed ITH. ITH may foster tumor evolution by providing diverse cell populations, and the dynamics of ITH architecture may evolve with neoplastic progression^8, 43^. Methylation ITH has been observed in various advanced malignancies and higher levels of methylation ITH have been reported to associate with inferior clinical outcomes^8, 44, 45, 46^. However, there are only a few reports on methylation ITH in precancers. For example, methylation ITH in Barrett esophagus, a precursor to esophageal ADC, was associated with the risk of malignant transformation^46^. In the current study, we demonstrated higher level of methylation ITH in later-stage IPNs than in early-stages (Fig. 3, Supplementary Fig. 4, 5). These results are in line with findings in advanced malignancies suggesting that complex methylation ITH may be associated with more aggressive tumor biology. As methylation ITH is linked to evolutionary plasticity and phenotypic diversity^44, 47, 48^, higher level of ITH could provide higher probability to survive and progress. Interestingly, eloci were significantly more abundant around TSSs in ADC than in AAH, AIS, or MIA. One plausible explanation is that although somatic methylation ITH may be stochastic during early lung carcinogenesis, some of the methylation aberrations (particularly those that are close to TSSs and potentially impact gene expression) may convey survival and/or growth advantages, resulting in selection of cells with higher densities of eloci around TSSs.

Parallel evolution of genome and methylome has been reported in various advanced malignancies including lung cancers^8, 49^. The current study demonstrated similar phylogenetic patterns and correlated genetic and methylation distances in IPNs of different stages (Fig. 5) suggesting genetic alterations and DNA methylation changes also evolve in parallel during early carcinogenesis of most lung ADCs. Meanwhile, promoter hypermethylation and copy number loss or mutations of putative TSGs were identified in independent IPNs within the same patients. These results were reminiscent of previous findings showing that distinct mutations of the same cancer genes were present in different regions of the same tumors^19, 50, 51^ or in different primary tumors from the same patients^52^, indicating convergent evolution. Although genetic events (e.g., copy number loss of TSGs) or methylation changes (e.g. promoter hypermethylation of TSGs) may independently or cooperatively offer proliferation or survival advantages to cells, these processes may be constrained around certain genes or pathways (e.g., inactivation of *TSC2* in the case of patient C10) that are essential to carcinogenesis in certain patients.

In our cohort, global hypomethylation was found to associate with significantly higher TMB, CNV burden as well as AI burden (Fig. 6b-d), consistent with previous reports that global hypomethylation is associated with CIN^32^ and increased rate of somatic mutations^33^. These data suggest that methylation aberrations have not only evolved in parallel with genomic aberrations but may have also facilitated accumulating genomic alterations that may have led to more drastic phenotypic changes in IPNs of later stages. Interestingly, higher-level of global hypomethylation was associated with higher proportion of clonal mutations (Fig. 6e). As the progression of lung precancers into invasive lung ADC predominantly follows a clonal sweeping model with selective outgrowth of fit subclones^5^, one plausible explanation for this association is that cells within the precancers with higher level of global hypomethylation may be prone to accumulate genomic aberrations, which subsequently provide growth advantages to these cell clones to develop into major clones in invasive lung ADC.

Cancer evolution results from accumulation of molecular alterations and is constantly shaped by selection pressure such as anti-tumor immune surveillance and therapeutic interventions. These molecular aberrations such as point mutations may accumulate gradually, a model termed microevolution, or in a punctuated or catastrophic manner through processes such as chromoplexy and chromothripsis, a model termed macroevolution, both of which have been reported in advanced malignancies^53, 54^. Largely due to lack of appropriate study materials, our understanding of the molecular evolutionary pattern during early carcinogenesis of lung ADC is rudimentary. Our previous study on genomic profiling of lung ADC and its precursors has demonstrated progressive accumulation of somatic mutations from AAH to AIS, MIA and ADC in line with the microevolution model. Meanwhile, there was a distinct increase in CNV burden from AAH to AIS and increase in AI burden from AIS to MIA. In the current study, we observed progressive increase of methylation changes from AAH to AIS, MIA and ADC. However, the overall difference in methylation aberrations appeared to be subtle between IPNs of different histologic stages (Fig.1b) and methylation evolution from AAH to ADC was similar across different epigenetically defined genomic regulatory regions. These observations suggest that methylation aberrations may have primarily contributed to the stepwise microevolution during early carcinogenesis of these lung ADCs and most methylation changes were stochastic “passengers”, as are the majority of somatic mutations^55^. On the other hand, these seemingly stochastic genomic and epigenetic alterations may give rise to heterogeneous subclones in precancers with various biological features, therefore increase the possibility of establishing fit subclones leading to malignant transformation. These principles may also apply to later neoplastic evolution including invasion, metastasis and development of drug resistance, where large-scale genomic sequencing studies have only depicted the underlying molecular mechanisms in a small subset of patients^56, 57^. Comprehensive molecular profiling incorporating genomic, epigenomics and transcriptomic profiling are warranted in future studies to depict these critical cancer evolutionary processes.

Anti-tumor immune surveillance plays a central role during initiation and progression of precancers. We have previously reported that the immune microenvironment was suppressed in invasive lung cancers compared to preinvasive cancers or precancers^58^. In this study, we demonstrated higher Treg/CD8 ratio in later stage IPNs (Fig. 7c, Supplementary Fig. 12c) implying a more suppressed T cell infiltrate in later-stage diseases, in line with a concomitant study of immune profiling of the same cohort of IPNs^59^. These findings are consistent with the concept of immune-editing, whereby the immunogenicity of cancer cells evolves under the selective pressure from anti-tumor immune response, resulting in the emergence of immune-resistant cancer clones in later stage diseases. Interestingly, global hypomethylation was associated with higher Treg/CD8 ratio (Fig. 7d). Methylation aberrations may affect anti-tumor immune surveillance directly by regulating the expression of immune-related genes^60^ and/or potential neoantigens or indirectly via modifying chromosomal vulnerability for CNV and mutations, both of which are well known to influence the tumor immune microenvironment^61, 62^. However, these impacts are complicated as many processes can affect anti-tumor immune surveillance both positively and negatively. For example, a high level of global hypomethylation may lead to a high CNV burden known to associate with a cold tumor immune microenvironment^61^; meanwhile global hypomethylation is also associated with increased mutation rate, which may increase tumor immunogenicity^62^. In the end, selection of cancer cell clones under immune pressure is determined by the cumulative effects of these molecular aberrations and only the cells with the best combination of molecular features including methylation status, mutation and CNV burden will survive and develop into dominant clones in invasive cancers.

There has been increasing enthusiasm toward moving interventions successfully applied to metastatic cancers to early stage cancers and even precancers, a concept called interception^63^. Compared with invasive cancers, precancers and preinvasive cancers may exhibit less complexity in aberrant molecular landscapes, as well as better preserved immune contextures, and thus may be easier to eradicate. Accordingly, we have launched the IMPRINT-Lung clinical trial (NCT03634241), in which patients with high-risk IPNs (many of which may be AAH or AIS) are treated with immune checkpoint inhibitors. In the current study, we demonstrated that DNA methylation aberrations are less complex in precancers and preinvasive lung cancers than in invasive cancers. Therefore, therapeutic agents that can modulate methylation by targeting aberrant methylations and potentially reprogram the immune microenvironment may also have potential in treating precancers and preinvasive cancers to prevent invasive lung cancers.

We delineated the evolution of genome-wide DNA methylation during the early carcinogenesis of lung ADC using RRBS on invasive lung ADC precursors of different stages. Rather than aggregated changes from populations of cancer cells measured by methylation array, RRBS assesses DNA methylation heterogeneity of single molecules derived from individual cancer cells, which made it possible to delineate methylation ITH at the allelic level. Furthermore, the WES data from the same IPNs has provided a unique opportunity to depict the relationship between methylation and genomic evolution. Due to the scarcity of IPN materials, our study has inevitable limitations. First, the sample size was relatively small for each histologic stage. Given the substantial heterogeneity between IPNs even within the same stages, our data was not powered to address some essential questions relevant to methylation evolution, such as whether certain methylation aberrations are significantly more common in early-stage IPNs than later-stage lesions representing “dead-end” IPNs. Second, most of these IPNs were very small after pathological assessment, which prevented acquisition of sufficient data in more patients for in-depth analyses such as multi-region profiling to dissect the interaction between parallel evolution and CIN; transcriptomic profiling to determine biological impact of observed methylation changes. Although a substantial number of DMRs were associated with TF binding sites, which could potentially regulate the transcriptome of the IPNs, these remained to be speculation without confirmation from transcriptomic data. Third, follow-up time for all patients in this study was relatively short, so we were unable to investigate the impact of these methylation changes on recurrence or survival. Finally, the resected specimens in this study could only provide a single molecular snapshot of the evolutionary process of IPNs. Whether all AAH will evolve into AIS, MIA, and ADC; whether all ADC evolve from AAH; and whether the observed methylation changes in IPNs of different stages represent the true evolutionary dynamics or simply reflect the distinct methylation patterns of IPNs with different malignant potentials is unknown. Deciphering the temporal evolution during neoplastic progression will require specimens obtained over the course of disease progression. Clinical trials collecting longitudinal biopsy specimens, such as IMPRINT-Lung (NCT03634241), may provide such opportunities going forward.

## METHODS

### Sample acquisition

A total of 53 resected pulmonary nodules and paired normal lung tissues from 39 patients treated at Nagasaki University Hospital or Zhejiang Cancer Hospital between 2014 and 2017 were used in the study. None of the patients received chemotherapy or radiotherapy before surgery. 29 lung nodules from 15 patients had multiregional specimens for spatial heterogeneity assessment (Supplementary Data 1). Whole-exome sequencing (WES) data was available for all specimens (EGAS00001004960)^5^.

### DNA methylation profiling by RRBS and data processing

DNA was extracted using the QIAamp DNA FFPE Tissue Kit (QIAGEN), and 200ng–1μg of DNA was subjected to RRBS for genome-wide DNA methylation profiling^64^. Briefly, TrimGalore v.0.4.3 was used to trim the Illumina adapter sequences (a minimum of 5 bp in a read was required to overlap with the adapter sequence); then Bismark v.0.18.1 integrating bowtie2 v.2.2.3 was used to align the trimmed reads to the GRCh37 assembly of the human genome. FastQC v.0.11.7 was used for quality control. The DNA methylation levels for individual CpGs were calculated using methyKit (v.1.16.0)^65^. Methylated reads (containing Cs) and unmethylated reads (containing Ts) at each cytosine site were counted, and the percentage of methylated reads among total reads covering the corresponding cytosine was calculated to quantify the DNA methylome for each sample at the single-base resolution. The CpG sites, DMRs, and loci of known genes, as well as genomic features, including CpG islands, were annotated using the R package “ChIPSeeker (v.1.26.0)”^66^ and the toolkit “genomation” (v.1.22.0)^67^, which is based on the “TxDb.Hsapiens.UCSC.hg19.knownGene” annotation database and the UCSC Genome Browser CpG islands table. To avoid bias, we kept CpG sites mapped to the autosome and removed CpG sites overlapping with the single-nucleotide polymorphism positions in dbSNP137. DNA methylation was analyzed at either a single-CpG resolution or at genomic region bins, in which DNA methylation values were averaged across 5-kb regions. Promoter methylation was calculated as the averaged DNA methylation values based on GENCODE promoter regions (i.e., 1 kb upstream to 500 bp downstream of the annotated TSS).

### Comparison of methylation profiles between different specimens

To assess the DNA methylome profiles of distinct IPNs of different pathological stages and examine the heterogeneity between IPN samples, we first aggregated the DNA methylation levels of 5-kb tiling regions across the genome in each sample by retaining only the CpG sites with ≥10 sequencing reads. We then applied principal component analysis to identify global DNA methylation patterns between samples. To evaluate consistent clusters of these DNA methylation profiles, we performed unsupervised hierarchical agglomerative analysis of CpG sites covered by ≥50 reads across all samples; these reads were based on single CpG methylation calls without any binning. To evaluate the correlation between overall DNA methylation in all samples from each patient, we used a pairwise approach to compare distance and similarity matrices on the basis for all CpGs with coverage of ≥10 reads.

### Differential DNA methylation analysis

Differentially methylated regions (DMRs) encompassing the differentially methylated CpGs between paired disease and normal tissue samples were identified using a triangular kernel to smooth the number of methylated reads and total number of reads by applying the “noise filter” function in the “DMR caller” (v.1.22.0) R package^68^. The differentially methylated CpG of paired samples were identified by selecting CpG sites located in DMRs; only CpG sites covered by ≥10 reads in paired samples were included.

### Partition of genomic regulatory regions based on chromatin states model

ChromHMM v.1.21 is applied to build the chromatin states model at 200 bp resolution with default parameters^12^ on A549 cell line (Encode Broad) for histone marks: H3K04me1, H3K04me2, H3K04me3, H3K09ac, H3K09me3, H3K27ac, H3K27me3, H3K36me3, and H3K79me2. The genomic regions including promoter regions, enhancer regions, transcribed regions and repressed regions (heterochromatin) were partitioned based on resulting segments inferred as defined chromatin states, which were deduced from chromatin-state signatures using a multivariate hidden Markov model (HMM) that explicitly models the combinatorial presence or absence of each histone mark.

### Repetitive elements

The repetitive elements were retrieved from UCSC repeatmasker track (https://genome.ucsc.edu/cgi-bin/hgTrackUi?g=rmsk), which was created using Arian Smit’s RepeatMasker program, through screening human genomic DNA sequences for interspersed repeats and low complexity DNA sequences.

### Identification of partially methylated domains (PMDs)

The boundary of partially methylated domains (PMDs) were inferred using whole genome shotgun bisulfite sequencing (WGSBS) of lung cells from a healthy donor (GSM983647) applying MethylSeekR (v.1.30.0) package^69^. Before PMD calling, CpGs overlapping common SNPs (dbSNP build 137) were removed.

### Motif identification and prediction of TF binding sites

The de novo methylated DNA motifs in IPNs of each stage were identified by mEpigram v.0.07, which discovers motifs by using position-specific weight matrices from the k-mers that are most enriched in the positive sequences compared with the negative sequences as “seeds” and extending the motifs in both directions^70^. The Tomtom tool (v.5.2.0) from the MEME suite was used to select significantly enriched methylated DNA motifs based on the database of known transcript factors (HOCOMOCO_v11)^71^.

### Estimation of DNA methylation ITH

To estimate DNA methylation ITH, we applied “methclone” (v.0.1) to identify epigenetic loci whose distributions of epigenetic allele (“epiallele”) clonality differed between paired tumor and normal samples by quantifying the degree to which the compositions of epialleles at given loci in the tumor samples were distinct from those in the normal tissue samples. An epiallele was defined by setting 60 reads in four consecutive CpG sites as the threshold to consider the epigenetic allele composition of the locus. We then calculated the differences in epiallele entropy between each IPN sample and its matched normal tissue sample. Loci with combinatorial entropy changes (*ΔS*) below –60 between each IPN sample and its paired normal sample were defined as epigenetic shift loci (termed “eloci”). To reduce the bias due to the different coverage for each sample, we then calculated relative epiallele shifts (i.e., the normalized number of eloci) via dividing the number of eloci by the total number of assessed loci in each sample and then multiplying that ratio by the average number of total loci across all samples^72^. We also assessed epigenetic polymorphism (“epipolymorphism”) to measure the epiallelic diversity in each IPN sample by calculating the frequency of each specific epiallele from multiple stochastic changes in the frequencies of many epialleles^73^. We calculated 16 epiallele status (0000, 0100, 0010, 0001, 1000, 1100, 0110, 0011, 0101, 1010, 1001,0111, 1011, 1101, 1110, and 1111, where 1 represents a methylated CpG site, and 0 represents an unmethylated CpG site), then epipolymorphism for each loci is defined as 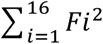,where *F* is the fraction of each epiallele status.

### Locus Overlap Analysis

We applied Locus Overlap Analysis (LOLA) ^23^ to the genomic regions identified as eloci (*ΔS*<–60) in all samples of each stage to evaluate the overlap between genomic regions with methylation ITH and chromatin marks. The genomic regions of all loci with ≥ 60 reads were used as background genomic regions, and the selected genomic regions were mapped to a compendium of publicly available histone mark profiles, including CTCF, H2AZ, H3K4me1, H3K4me2, H3K4me3, H3K9ac, H3K9me3, H3K27me3, H3K27ac, H3K36me3, H3K79me2, H4K20me1 in the A549 lung adenocarcinoma cell line (http://hgdownload.soe.ucsc.edu/goldenPath/hg19/encodeDCC/wgEncodeBroadHistone)and H3K27me3, H3K4me3, H3K9me2, H3K9ac, CTCF in an immortalized human bronchial epithelial cell line (BEAS-2B; GSE56053)^25^. P-values in the enrichment analyses were calculated using one-sided Fisher’s exact test. Adjustment for multiple testing with q-value (FDR adjusted p-value) was performed using the Benjamini–Yekutieli method.

### Construction of phylogenetic trees

To construct methylation-based phylogenetic trees from the RRBS data, we used promoter CpG sites (≥50 reads per CpG site selected for all samples from each patient) with the most variable methylation values (mean absolute deviation >10%) shared by all samples, including a normal tissue sample used as the tree root, to build a Euclidean distance matrix. We built the phylogenetic trees by applying a neighbor-joining algorithm from the “ape” (v.5.4.1) package to independently infer phylogenetic relationships between IPN specimens for each patient from the mutation and methylation profiles. To assess the phylogenetic similarity between the genetic and epigenetic profiles, we employed an independent but parallel distance matrix construction from the mutation and methylation profiles for each IPN, in which the genetic distance was quantified by Hamming distance based on all filtered mutations and methylation distance was quantified by Euclidean distance based on methylation values of all CpG sites (≥20 reads, MAD≥20 for all samples in each IPN), then calculated the Spearman correlation coefficient for all pairwise samples grouped by IPNs.

### Estimation of global hypomethylation

To determine global methylation levels, we chose CpG sites (covered by ≥10 aligned reads) within evolutionarily young subfamilies of LINE-1 repeat elements (L1HS and L1PA). LINE-1 family annotation were obtained from the Repeat-Masker of the UCSC genome browser. We averaged the methylation values of the chosen CpG sites, to represent the global methylation level of each sample.

### Deconvolution of T cell profiles

To derive tumor-infiltrating T cell subtypes from transcriptomic data previously published^36^, processed RNAseq dataset (normalized and log2 transformed) comprise of 197 normal lung tissues, 98 AIS/MIA and 99 ADC samples were retrieved from EGAS00001004006. ImmuCellAI^74^ was applied to infer immune cell components for each sample.

To derive the infiltration of T cell repertoire from RRBS data, we first obtained a reference methylation signature by retrieving the methylation calls of whole-genome bisulfite datasets (WGBS) from the BLUEPRINT epigenome project, including those for samples of regulatory T cells (EGAX00001343016/EGAX00001236257 in EGAD00001002492) and CD8+ T cells (EGAX00001195937/EGAX00001195943 in EGAD00001002486).Only promoter CpG sites that were covered by ≥5 reads in all samples were retained and binned with a 50-bp window by mean methylation values, then only 50-bp binned regions that overlapped with 50-bp binned regions of CpG sites covered by the RRBS methylation profiles of preneoplastic lesions in all samples were used for reference signature extraction by non-negative matrix factorization (NMF) implemented in the “MethylCIBERSORT” (v.0.2.0) package. We then performed DNA methylation deconvolution (average methylation level by 50-bp bin) using the aforementioned BLUEPRINT signature and the CIBERSORT webserver (https://cibersort.stanford.edu). We performed T cell deconvolution in relative mode, running 100 permutations with quantile normalization disabled^75^. The resulting immune-cellular fractions were used to compare samples of different pathological stages. The average values of immune cell infiltration for each sample inferred by whole-genome bisulfite analysis from two patients independently were used to infer Treg/CD8 ratio.

### Statistical analysis

Violin plots were created using the “geom_violin” function in R statistical package ggplot2 (v0.9.1) to represent data point density along the *Y*-axis, and the “stat_summary” function from ggplot2 (v0.9.1) was used to calculate the mean as the center point. Differences in DMR numbers, eloci numbers, and immune cell infiltration between IPN specimens of different stages were assessed using the Kruskal–Wallis chi-square test. We used quantile regression to assess the differential entropy values at 1th percentile (low percentile of entropy was chosen to reflect the loci with biological interest) and epipolymorphism values at 50th percentile between different groups. We used a two-sided Pearson correlation coefficient to compare methylation profiles between two samples and between groups of samples of different stages. We used a two-sided Spearman correlation coefficient to determine the extent to which distance matrices were correlated with DNA methylation profiles and somatic mutation profiles.

### Data availability

The RRBS dataset has been deposited at European Bioinformatics Institute European Genome–phenome Archive (EGA) (accession number: EGAS00001004610, https://www.ebi.ac.uk/ega/studies/EGAS00001004610) through controlled access. Whole-exome sequencing is under the accession code: EGAD00001004960 [https://www.ebi.ac.uk/ega/datasets/EGAD00001004960]. To protect patient privacy, interested researchers need to apply via data access committee (DAC), which will grant all reasonable requests. GSM983647 (Whole Genome Shotgun Bisulfite Sequencing of human normal lung cells) was downloaded from GEO [https://www.ncbi.nlm.nih.gov/geo/query/acc.cgi?acc=GSM983647].

EGAS00001004006 (RNA-seq of human preinvasive and invasive lung adenocarcinoma) was downloaded from EGA [https://www.ebi.ac.uk/ega/studies/EGAS00001004006].

EGAD00001002492 (Bisulfite-seq of regulatory T cells from human venous blood) [https://www.ebi.ac.uk/ega/datasets/EGAD00001002492] and EGAD00001002486 (Bisulfite-seq of CD8+ T cells from human venous blood) [https://www.ebi.ac.uk/ega/datasets/EGAD00001002486] was downloaded from EGA, respectively. And source data are provided with this study. All other data may be found within the main manuscript or supplementary information or available from the authors upon request.

## Data Availability

FASTQ files for RRBS dataset is uploading to EGA and available upon request.

https://www.ebi.ac.uk/ega/studies/EGAS00001004610

## AUTHOR CONTRIBUTIONS

X.H. and J.J.Z. conceived the study and wrote the manuscript. J.J.Z. and P.A.F. jointly supervised and financially supported the study. X.H. performed all bioinformatics and statistical data analyses in consultation with M.R.E., J.P.I., L.H.W., J.J.L., and Y.L.; J.F.J. supervised pathological assessments and the preparation of specimens. L.S.Y., J.F.K., and H.P. collected resected specimens and clinical data. C.W.C., C.B, and R.Z.C. prepared DNA samples. M.C.G. and B.W.C. performed radiological assessment. M.R.E. supervised RRBS profiling. X.H., M.R.E., R.Z.C., A.R., N.M., L.H.W., J.C.Z., J.H.Z., P.S., A.A.V., J.V.H., I.I.W., H.K., M.B.A, B.S., H.P., J.P.I., D.S., P.A.F., and J.J.Z. interpreted the data. All authors reviewed the manuscript.

## ACKNOWLEDGMENTS

This study was supported by the MD Anderson Khalifa Scholar Award, the National Cancer Institute of the National Institute of Health Research Project Grant (R01CA234629-01), the AACR-Johnson & Johnson Lung Cancer Innovation Science Grant (18-90-52-ZHAN), the MD Anderson Physician Scientist Program, the MD Anderson Lung Cancer Moon Shot Program, Sabin Family Foundation Award, Duncan Family Institute Cancer Prevention Research Seed Funding Program, The University of Texas MD Anderson Cancer Center Pre-Cancer Atlas Project, EDRN U01 (U01CA214195), the Cancer Prevention and Research Institute of Texas Multi-Investigator Research Award grant (RP160668). We thank MD Anderson Cancer Center’s Epigenomics Profiling Core and its Science Park Next-Generation Sequencing Core (supported by CPRIT Core Facility Support Award #RP120348) for performing RRBS profiling. We thank Sally Boyd, Jinzhen Chen, and Rong Yao, Stan Bujnowski, Eric Sisson for providing excellent technical support for high-performance cluster resource; we thank Joe Munch from Scientific Publications in MD Anderson’s Research Medical Library for editing the manuscript.

## References

1. Kadara H, Scheet P, Wistuba, II, Spira AE. Early Events in the Molecular Pathogenesis of Lung Cancer. Cancer Prev Res (Phila) 9, 518–527 (2016).

2. Chen Z, Fillmore CM, Hammerman PS, Kim CF, Wong KK. Non-small-cell lung cancers: a heterogeneous set of diseases. Nat Rev Cancer 14, 535–546 (2014).

3. Lee HJ, et al. IASLC/ATS/ERS International Multidisciplinary Classification of Lung Adenocarcinoma: novel concepts and radiologic implications. J Thorac Imaging 27, 340–353 (2012).

4. Chen D, Dai C, Kadeer X, Mao R, Chen Y, Chen C. New horizons in surgical treatment of ground-glass nodules of the lung: experience and controversies. Ther Clin Risk Manag 14, 203–211 (2018).

5. Hu X, et al. Multi-region exome sequencing reveals genomic evolution from preneoplasia to lung adenocarcinoma. Nat Commun 10, 2978 (2019).

6. Jones PA, Baylin SB. The epigenomics of cancer. Cell 128, 683–692 (2007).

7. Widschwendter M, et al. Epigenetic stem cell signature in cancer. Nat Genet 39, 157–158 (2007).

8. Quek K, et al. DNA methylation intratumor heterogeneity in localized lung adenocarcinomas. Oncotarget 8, 21994–22002 (2017).

9. Kerr KM, Galler JS, Hagen JA, Laird PW, Laird-Offringa IA. The role of DNA methylation in the development and progression of lung adenocarcinoma. Dis Markers 23, 5–30 (2007).

10. Selamat SA, et al. DNA methylation changes in atypical adenomatous hyperplasia, adenocarcinoma in situ, and lung adenocarcinoma. PLoS One 6, e21443 (2011).

11. Carter SL, et al. Absolute quantification of somatic DNA alterations in human cancer. Nat Biotechnol 30, 413–421 (2012).

12. Ernst J, Kellis M. Chromatin-state discovery and genome annotation with ChromHMM. Nat Protoc 12, 2478–2492 (2017).

13. Salhab A, et al. A comprehensive analysis of 195 DNA methylomes reveals shared and cell-specific features of partially methylated domains. Genome Biol 19, 150 (2018).

14. Yin Y, et al. Impact of cytosine methylation on DNA binding specificities of human transcription factors. Science 356, (2017).

15. Casey SC, Baylot V, Felsher DW. The MYC oncogene is a global regulator of the immune response. Blood 131, 2007–2015 (2018).

16. Wils LJ, Bijlsma MF. Epigenetic regulation of the Hedgehog and Wnt pathways in cancer. Crit Rev Oncol Hematol 121, 23–44 (2018).

17. Yu T, et al. KLF4 regulates adult lung tumor-initiating cells and represses K-Ras-mediated lung cancer. Cell Death Differ 23, 207–215 (2016).

18. Wu C, et al. WT1 promotes invasion of NSCLC via suppression of CDH1. J Thorac Oncol 8, 1163–1169 (2013).

19. Zhang J, et al. Intratumor heterogeneity in localized lung adenocarcinomas delineated by multiregion sequencing. Science 346, 256–259 (2014).

20. Reuben A, et al. Comprehensive T cell repertoire characterization of non-small cell lung cancer. Nat Commun 11, 603 (2020).

21. Gaidatzis D, et al. DNA sequence explains seemingly disordered methylation levels in partially methylated domains of Mammalian genomes. PLoS Genet 10, e1004143 (2014).

22. Lochs SJA, Kefalopoulou S, Kind J. Lamina Associated Domains and Gene Regulation in Development and Cancer. Cells 8, (2019).

23. Sheffield NC, Bock C. LOLA: enrichment analysis for genomic region sets and regulatory elements in R and Bioconductor. Bioinformatics 32, 587–589 (2016).

24. Consortium EP. An integrated encyclopedia of DNA elements in the human genome. Nature 489, 57–74 (2012).

25. Jose CC, et al. Epigenetic dysregulation by nickel through repressive chromatin domain disruption. Proc Natl Acad Sci U S A 111, 14631–14636 (2014).

26. Lienert F, et al. Genomic prevalence of heterochromatic H3K9me2 and transcription do not discriminate pluripotent from terminally differentiated cells. PLoS Genet 7, e1002090 (2011).

27. Kondo Y, et al. Gene silencing in cancer by histone H3 lysine 27 trimethylation independent of promoter DNA methylation. Nat Genet 40, 741–750 (2008).

28. Hon GC, et al. Global DNA hypomethylation coupled to repressive chromatin domain formation and gene silencing in breast cancer. Genome Res 22, 246–258 (2012).

29. Ohm JE, et al. A stem cell-like chromatin pattern may predispose tumor suppressor genes to DNA hypermethylation and heritable silencing. Nat Genet 39, 237–242 (2007).

30. Schlesinger Y, et al. Polycomb-mediated methylation on Lys27 of histone H3 pre-marks genes for de novo methylation in cancer. Nat Genet 39, 232–236 (2007).

31. Astrinidis A, Cash TP, Hunter DS, Walker CL, Chernoff J, Henske EP. Tuberin, the tuberous sclerosis complex 2 tumor suppressor gene product, regulates Rho activation, cell adhesion and migration. Oncogene 21, 8470–8476 (2002).

32. Eden A, Gaudet F, Waghmare A, Jaenisch R. Chromosomal instability and tumors promoted by DNA hypomethylation. Science 300, 455 (2003).

33. Chen RZ, Pettersson U, Beard C, Jackson-Grusby L, Jaenisch R. DNA hypomethylation leads to elevated mutation rates. Nature 395, 89–93 (1998).

34. Saito K, Kawakami K, Matsumoto I, Oda M, Watanabe G, Minamoto T. Long interspersed nuclear element 1 hypomethylation is a marker of poor prognosis in stage IA non-small cell lung cancer. Clin Cancer Res 16, 2418–2426 (2010).

35. Reuben A, et al. TCR Repertoire Intratumor Heterogeneity in Localized Lung Adenocarcinomas: An Association with Predicted Neoantigen Heterogeneity and Postsurgical Recurrence. Cancer Discov 7, 1088–1097 (2017).

36. Chen H, et al. Genomic and immune profiling of pre-invasive lung adenocarcinoma. Nat Commun 10, 5472 (2019).

37. Chakravarthy A, et al. Pan-cancer deconvolution of tumour composition using DNA methylation. Nat Commun 9, 3220 (2018).

38. Peng GL, et al. CD8(+) cytotoxic and FoxP3(+) regulatory T lymphocytes serve as prognostic factors in breast cancer. Am J Transl Res 11, 5039–5053 (2019).

39. Chen RZ, et al. T cell repertoire evolution from the normal lung to invasive lung adenocarcinoma. Cancer Research 78, (2018).

40. Cancer Genome Atlas Research N. Comprehensive molecular profiling of lung adenocarcinoma. Nature 511, 543–550 (2014).

41. Saghafinia S, Mina M, Riggi N, Hanahan D, Ciriello G. Pan-Cancer Landscape of Aberrant DNA Methylation across Human Tumors. Cell Rep 25, 1066–1080 e1068 (2018).

42. Chakravarthy A, et al. Pan-cancer deconvolution of tumour composition using DNA methylation. Nature Communications 9, (2018).

43. Prasetyanti PR, Medema JP. Intra-tumor heterogeneity from a cancer stem cell perspective. Mol Cancer 16, 41 (2017).

44. Landau DA, et al. Locally disordered methylation forms the basis of intratumor methylome variation in chronic lymphocytic leukemia. Cancer Cell 26, 813–825 (2014).

45. Hua X, et al. Genetic and epigenetic intratumor heterogeneity impacts prognosis of lung adenocarcinoma. Nat Commun 11, 2459 (2020).

46. Merlo LM, et al. A comprehensive survey of clonal diversity measures in Barrett’s esophagus as biomarkers of progression to esophageal adenocarcinoma. Cancer Prev Res (Phila) 3, 1388–1397 (2010).

47. Siegmund KD, Marjoram P, Woo YJ, Tavare S, Shibata D. Inferring clonal expansion and cancer stem cell dynamics from DNA methylation patterns in colorectal cancers. Proc Natl Acad Sci U S A 106, 4828–4833 (2009).

48. Hansen KD, et al. Increased methylation variation in epigenetic domains across cancer types. Nat Genet 43, 768–775 (2011).

49. Aryee MJ, et al. DNA methylation alterations exhibit intraindividual stability and interindividual heterogeneity in prostate cancer metastases. Sci Transl Med 5, 169ra110 (2013).

50. Jamal-Hanjani M, et al. Tracking the Evolution of Non-Small-Cell Lung Cancer. N Engl J Med 376, 2109–2121 (2017).

51. consortium TRR. TRACERx Renal: tracking renal cancer evolution through therapy. Nat Rev Urol 14, 575–576 (2017).

52. Liu Y, et al. Genomic heterogeneity of multiple synchronous lung cancer. Nat Commun 7, 13200 (2016).

53. Shen MM. Chromoplexy: a new category of complex rearrangements in the cancer genome. Cancer Cell 23, 567–569 (2013).

54. Cortes-Ciriano I, et al. Comprehensive analysis of chromothripsis in 2,658 human cancers using whole-genome sequencing. Nat Genet 52, 331–341 (2020).

55. Flavahan WA, Gaskell E, Bernstein BE. Epigenetic plasticity and the hallmarks of cancer. Science 357, (2017).

56. Zheng JB, et al. DNA methylation affects metastasis of renal cancer and is associated with TGF-beta/RUNX3 inhibition. Cancer Cell International 18, (2018).

57. Le X, et al. Landscape of EGFR-Dependent and -Independent Resistance Mechanisms to Osimertinib and Continuation Therapy Beyond Progression in EGFR-Mutant NSCLC. Clin Cancer Res 24, 6195–6203 (2018).

58. Sivakumar S, et al. Genomic Landscape of Atypical Adenomatous Hyperplasia Reveals Divergent Modes to Lung Adenocarcinoma. Cancer Res 77, 6119–6130 (2017).

59. Dejima H, et al. Immune evolution from preneoplasia to invasive lung adenocarcinomas and underlying molecular features. medRxiv, 2020.2007.2011.20142992 (2020).

60. Liu M, Zhou J, Chen Z, Cheng AS. Understanding the epigenetic regulation of tumours and their microenvironments: opportunities and problems for epigenetic therapy. J Pathol 241, 10–24 (2017).

61. Davoli T, Uno H, Wooten EC, Elledge SJ. Tumor aneuploidy correlates with markers of immune evasion and with reduced response to immunotherapy. Science 355, (2017).

62. Jung H, et al. DNA methylation loss promotes immune evasion of tumours with high mutation and copy number load. Nat Commun 10, 4278 (2019).

63. Blackburn EH. Cancer interception. Cancer Prev Res (Phila) 4, 787–792 (2011).

64. Gu H, Smith ZD, Bock C, Boyle P, Gnirke A, Meissner A. Preparation of reduced representation bisulfite sequencing libraries for genome-scale DNA methylation profiling. Nat Protoc 6, 468–481 (2011).

65. Akalin A, et al. methylKit: a comprehensive R package for the analysis of genome-wide DNA methylation profiles. Genome Biol 13, R87 (2012).

66. Yu G, Wang LG, He QY. ChIPseeker: an R/Bioconductor package for ChIP peak annotation, comparison and visualization. Bioinformatics 31, 2382–2383 (2015).

67. Akalin A, Franke V, Vlahovicek K, Mason CE, Schubeler D. Genomation: a toolkit to summarize, annotate and visualize genomic intervals. Bioinformatics 31, 1127–1129 (2015).

68. Catoni M, Tsang JM, Greco AP, Zabet NR. DMRcaller: a versatile R/Bioconductor package for detection and visualization of differentially methylated regions in CpG and non-CpG contexts. Nucleic Acids Res 46, e114 (2018).

69. Burger L, Gaidatzis D, Schubeler D, Stadler MB. Identification of active regulatory regions from DNA methylation data. Nucleic Acids Res 41, e155 (2013).

70. Ngo V, Wang M, Wang W. Finding de novo methylated DNA motifs. Bioinformatics 35, 3287–3293 (2019).

71. Bailey TL, et al. MEME SUITE: tools for motif discovery and searching. Nucleic Acids Res 37, W202–208 (2009).

72. Li S, et al. Dynamic evolution of clonal epialleles revealed by methclone. Genome Biol 15, 472 (2014).

73. Landan G, et al. Epigenetic polymorphism and the stochastic formation of differentially methylated regions in normal and cancerous tissues. Nat Genet 44, 1207–1214 (2012).

74. Miao YR, et al. ImmuCellAI: A Unique Method for Comprehensive T-Cell Subsets Abundance Prediction and its Application in Cancer Immunotherapy. Adv Sci (Weinh) 7, 1902880 (2020).

75. Chen B, Khodadoust MS, Liu CL, Newman AM, Alizadeh AA. Profiling Tumor Infiltrating Immune Cells with CIBERSORT. Methods Mol Biol 1711, 243–259 (2018).

