## Supplementary Figure 1-12 for "Evolution of DNA methylome from precancerous lesions to invasive lung adenocarcinomas"

Hu, et al.

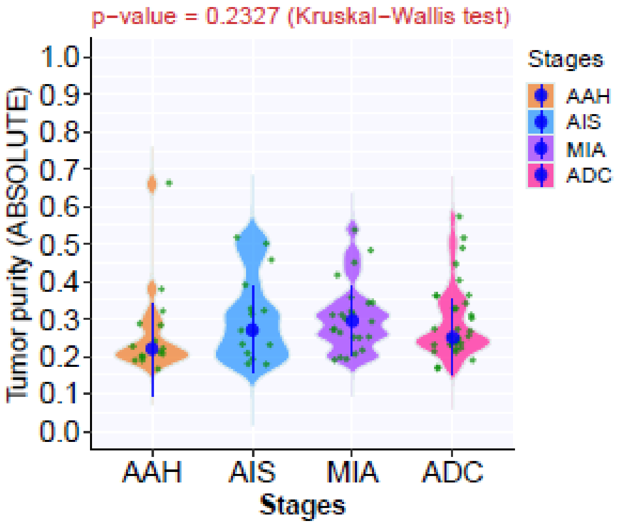


Supplementary Figure 1. The tumor purity in IPNs of different histologic stages. The tumor purity was derived from whole exome sequencing data using ABSOLUTE. The green dots represent tumor purity in each sample. The solid blue dots represent the median tumor purity in IPNs of each histologic stage with 95% confidence interval as error bars. Differences among different stages were assessed using the Kruskal-Wallis H test.

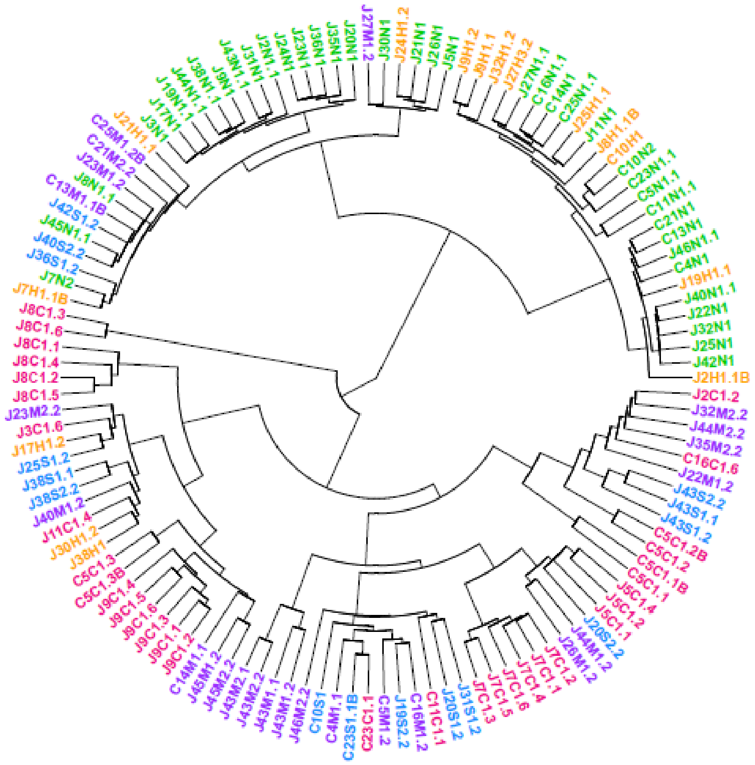

Supplementary Figure 2. Unsupervised hierarchical clustering of promoter DNA methylation. Unsupervised hierarchical clustering was performed using 2,261 CpG sites shared across all samples with minimum coverage of 50 reads per CpG site with a median absolute deviation (MAD) of >50. The colors of the sample names denote samples originating from normal tissue (green), AAH (orange), AIS (blue), MIA (purple), and ADC (rose).

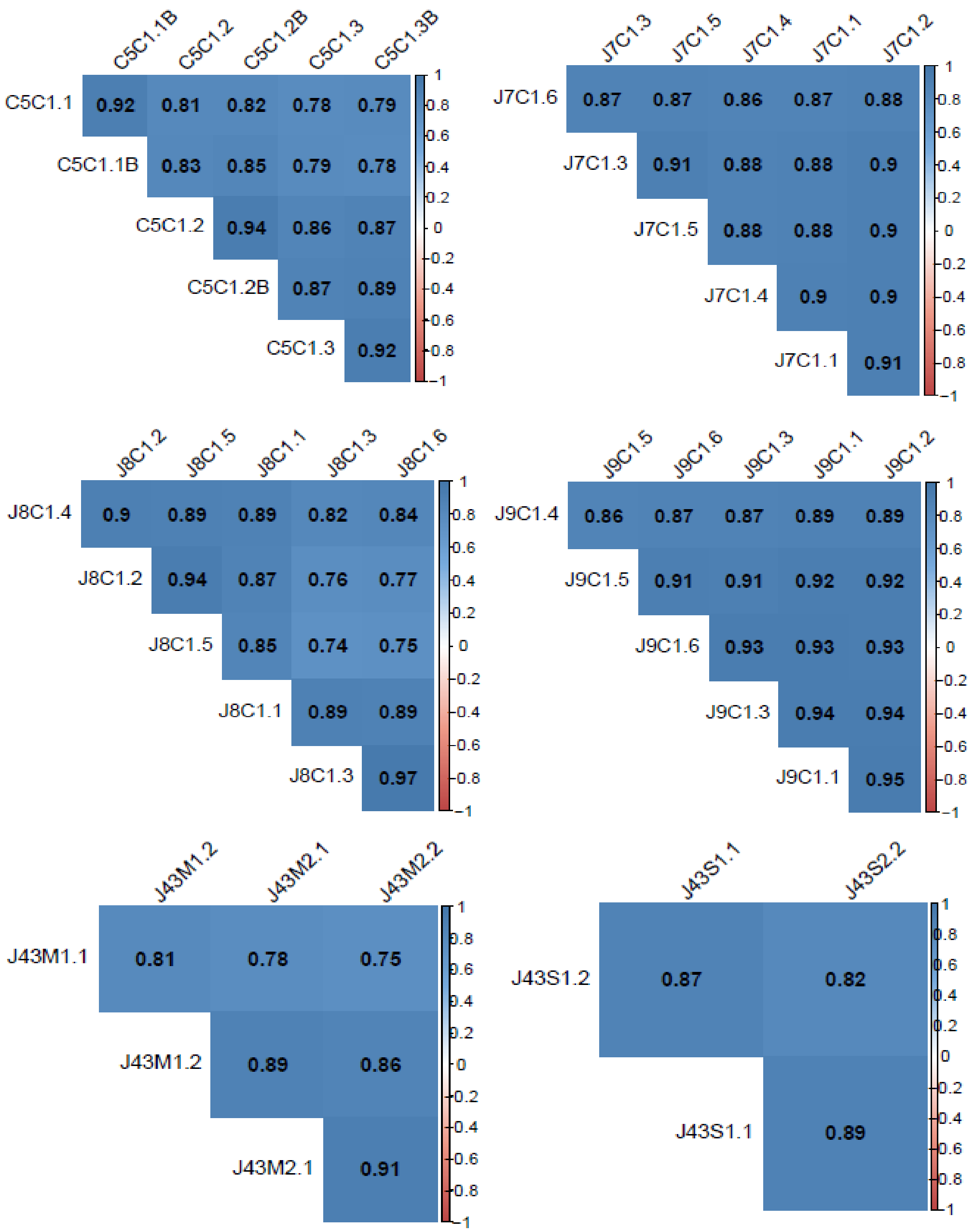


Supplementary Figure 3. Comparison of global DNA methylation patterns between different regions from the same IPNs. The heat-maps display the two-tailed Pearson correlation coefficients for pairwise comparisons of all samples from each IPN.

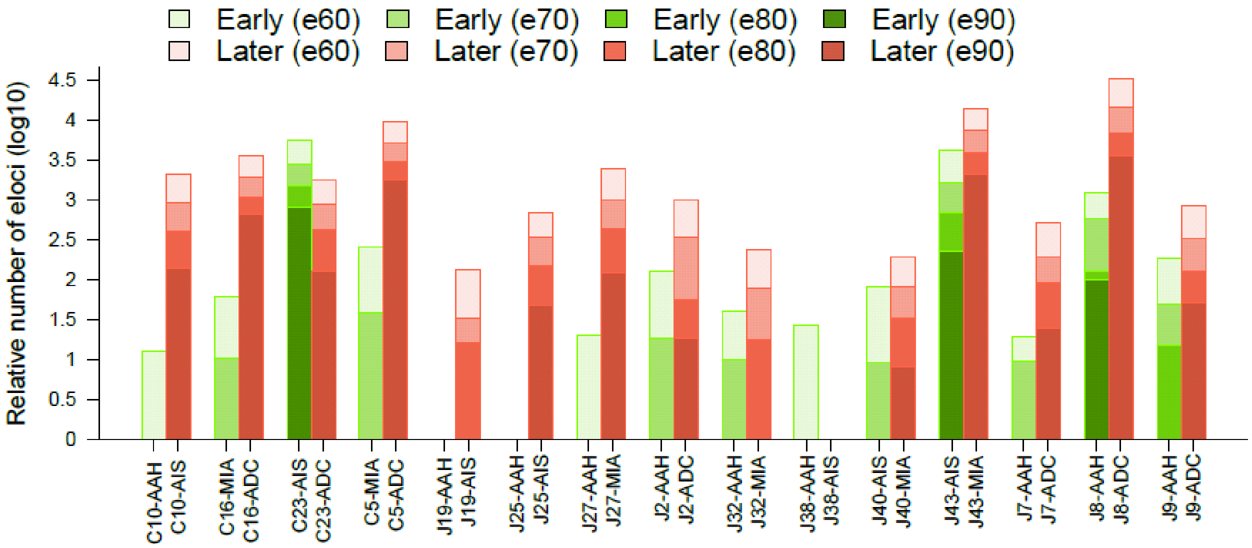


Supplementary Figure 4. Comparison of relative numbers of eloci in IPNs of different stages within the same patients. Each bar represents the relative number (log10-transformed) of eloci composed of 4 adjacent CpG sites for each IPN. Pairwise comparison of an early-stage IPN and a later-stage IPN from the same patient was performed using four different combinatorial entropy difference cutoffs (e90: -90, e80: -80, e70: -70, and e60: -60). Early stages: green, later stage: brown.

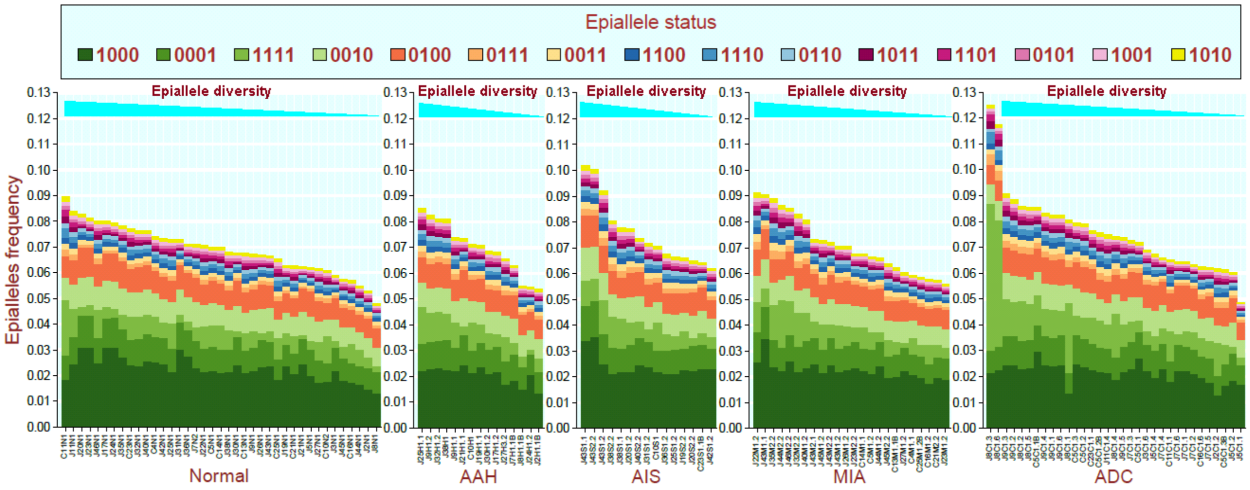


Supplementary Figure 5. The distribution of epigenetic status. The epiallele frequencies across autosomal regions for all IPN specimens of each histologic stage. Different combinations of four consecutive CpG sites at epiallele loci are color-coded. The 0s indicate unmethylated CpG sites and the 1s indicate methylated CpG sites. The “0000”, accounting for ~80%-90% of epialleles is not displayed.

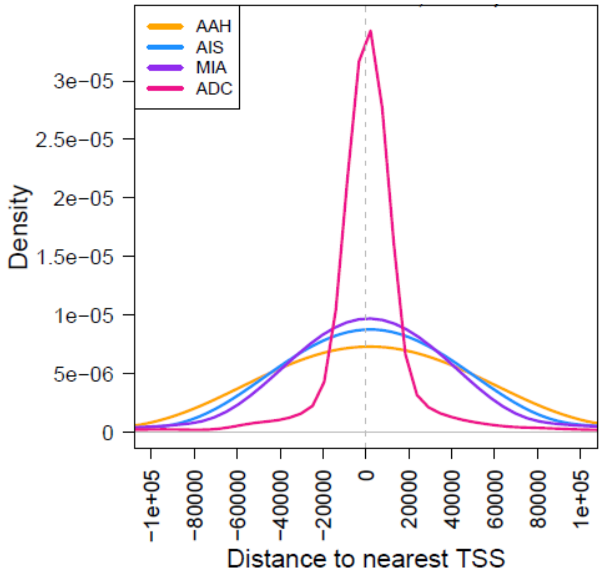


Supplementary Figure 6. The distances of eloci to nearest transcription start sites (TSSs) in IPNs of different stages. The density plot shows the distances of eloci ($\Delta S$<-60) to the nearest TSSs all AAH, AIS, MIA, and ADC specimens.

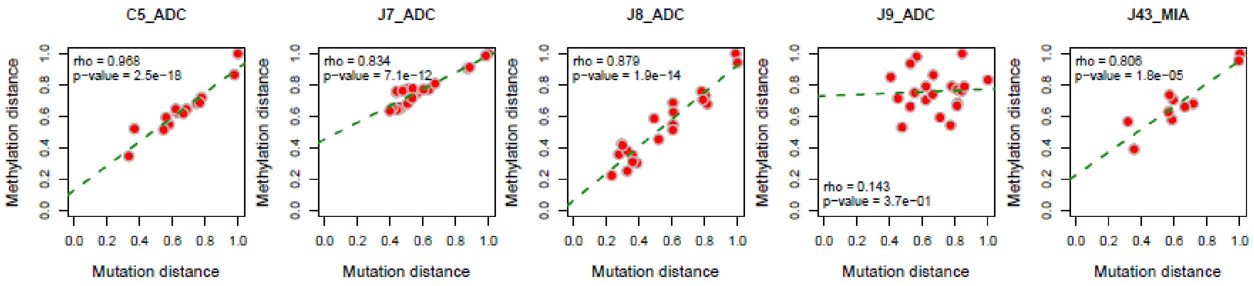


Supplementary Figure 7. Correlation of genetic distance and methylation distance between spatially separated regions from the same IPNs. The genetic distance was quantified by Hamming distance based on all mutations and methylation distance was quantified by Euclidean distance based on methylation values of all CpG sites. The correlation of genetic distance and methylation distance was assessed by two-tailed Spearman's correlation test. Each dot represents the normalized distance between each pair of specimens from the same IPN. Only IPNs with more than 4 spatially separated regions were included.

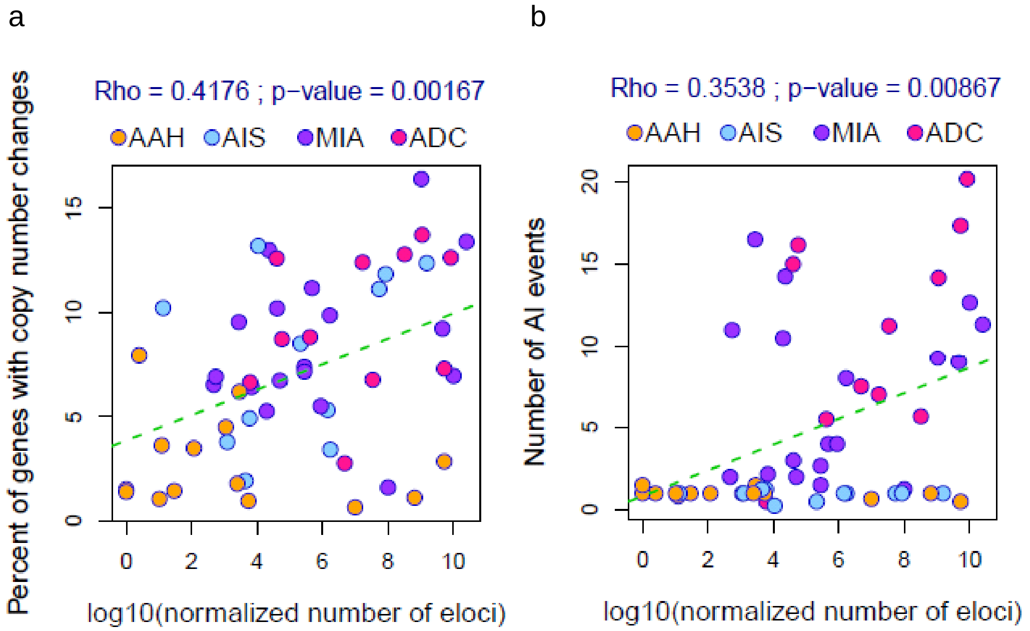


Supplementary Figure 8. Correlation of methylation ITH with genomic instability in IPNs of different stages. The relative number of eloci (*X*-axis) was used as a surrogate for methylation ITH and percent of genes with copy number changes (a), number of events with allelic imbalance (AI) (b) (*Y*-axis) were used as surrogates for chromosomal instability. The correlation was assessed by two-tailed Spearman's correlation test. Each dot represents each IPN specimen.

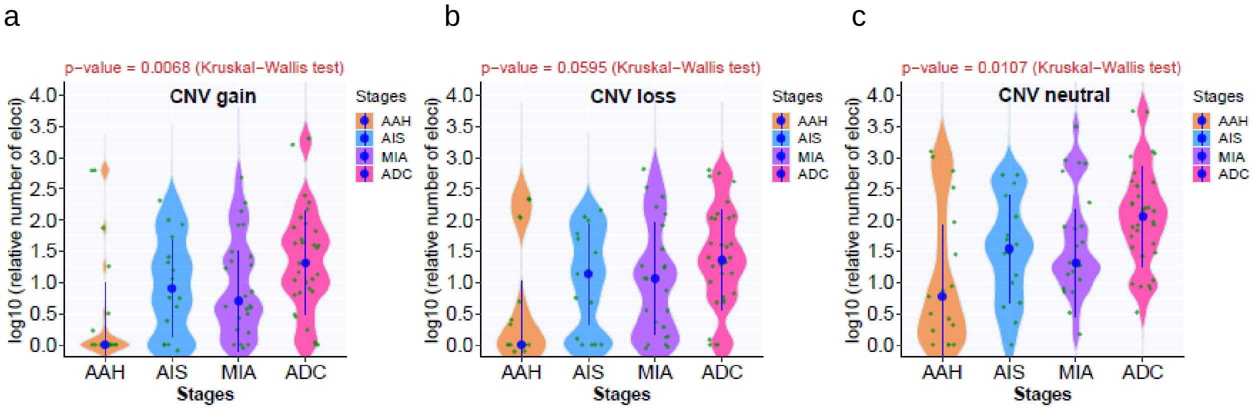


Supplementary Figure 9. Loci with methylation ITH located in genomic regions of different CNV status. The numbers of eloci in IPNs of different stages based on corresponding CpG sites located in genomic regions with copy number gain (a), loss (b) or neutral (c). The green dots represent normalized numbers of eloci in each IPN and the blue dots represent the mean of normalized numbers of eloci in IPNs of each histologic stage with 95% confidence interval as error bars. The differences among all stages were assessed using the Kruskal-Wallis H test.

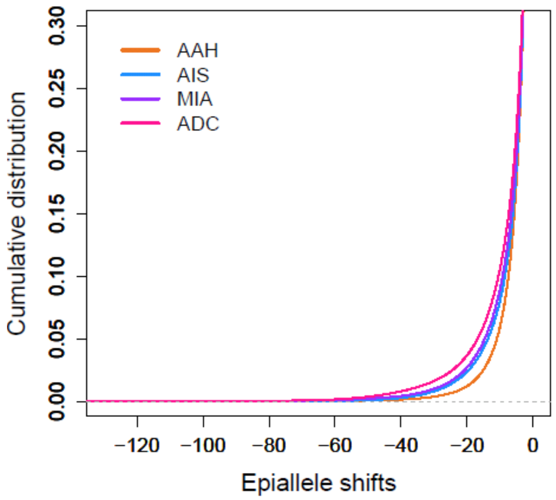


Supplementary Figure 10. Increased cumulative distribution of epiallele shifts in later-stage IPNs using loci located in copy number-neutral chromosomal regions. Cumulative distribution curves of epiallele shifts of the DNA methylome in AAH, AIS, MIA, and ADC specimens compared to normal lung, with 1th percentile of entropy as -21.52 for AAH, -27.96 for AIS, -30.03 for MIA, -36.84 for ADC by quantile regression.

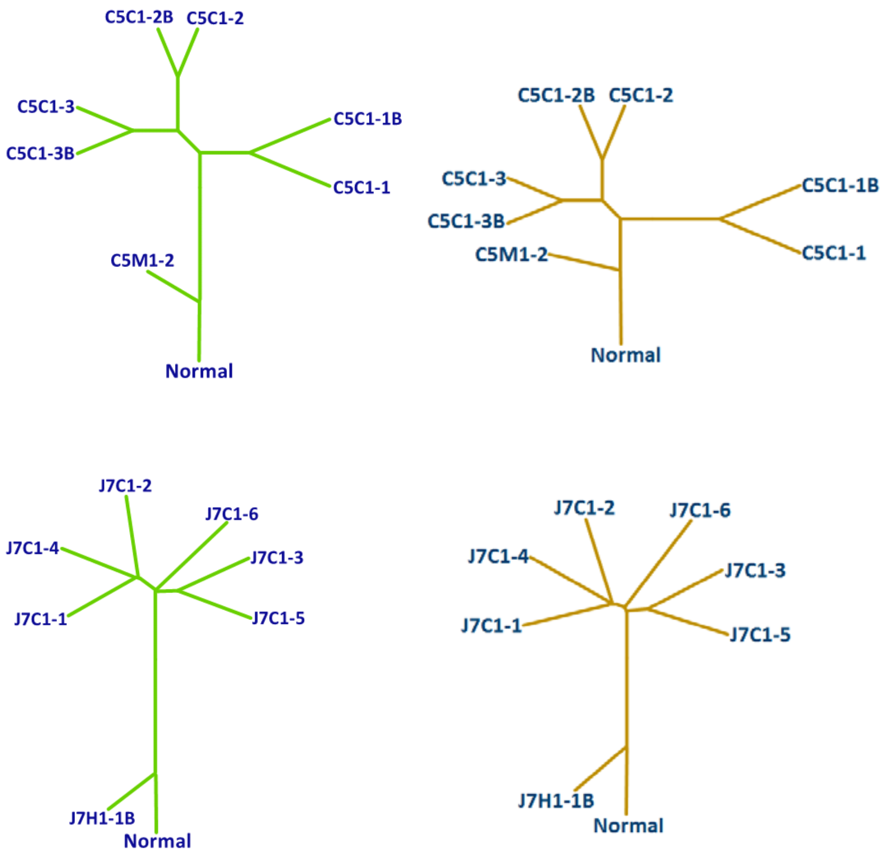

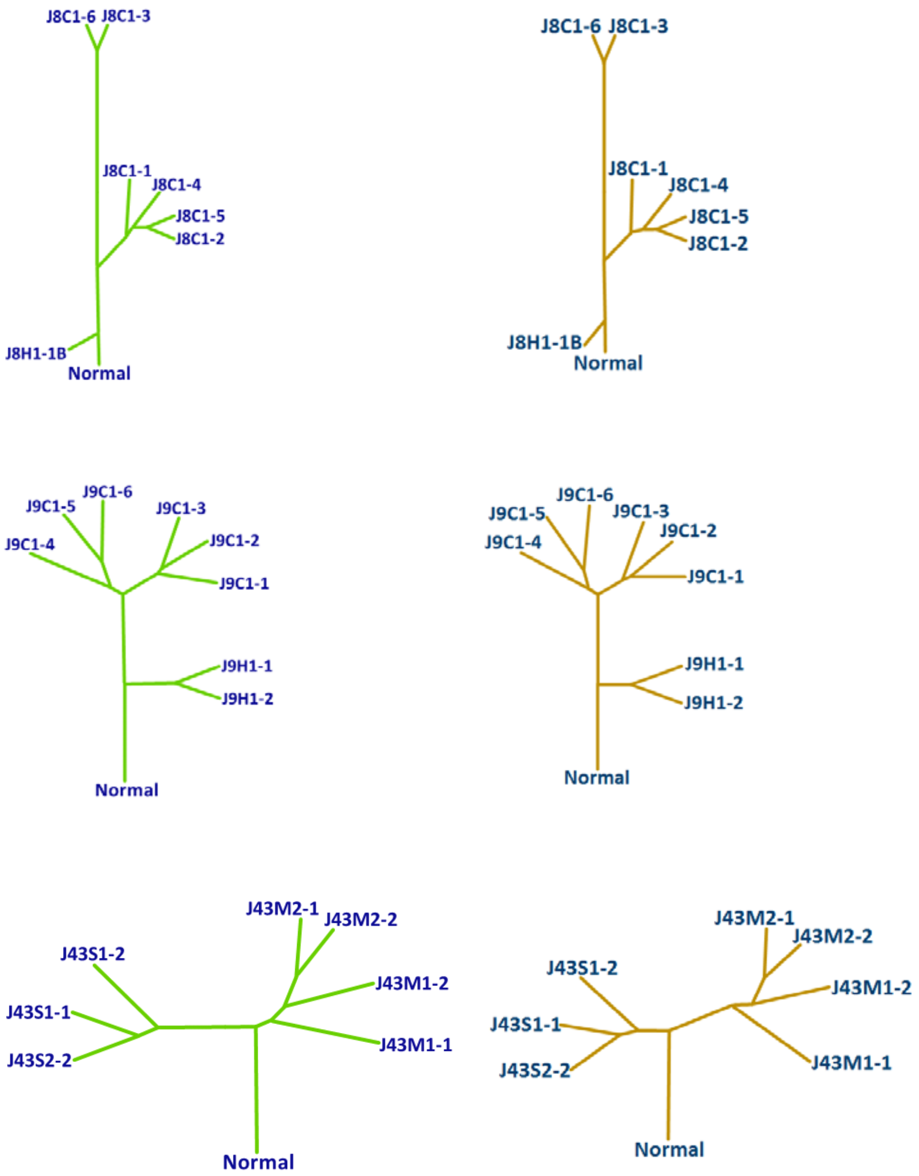


Supplementary Figure 11. Phylogenetic trees using all loci (left) versus using only loci located in copy number-neutral chromosomal regions (right). The length of each branch is proportional to the methylation distance between any pair of two spatially separated specimens from the same patients. Only patients with IPNs having at least 4 regional samples were included in this analysis.

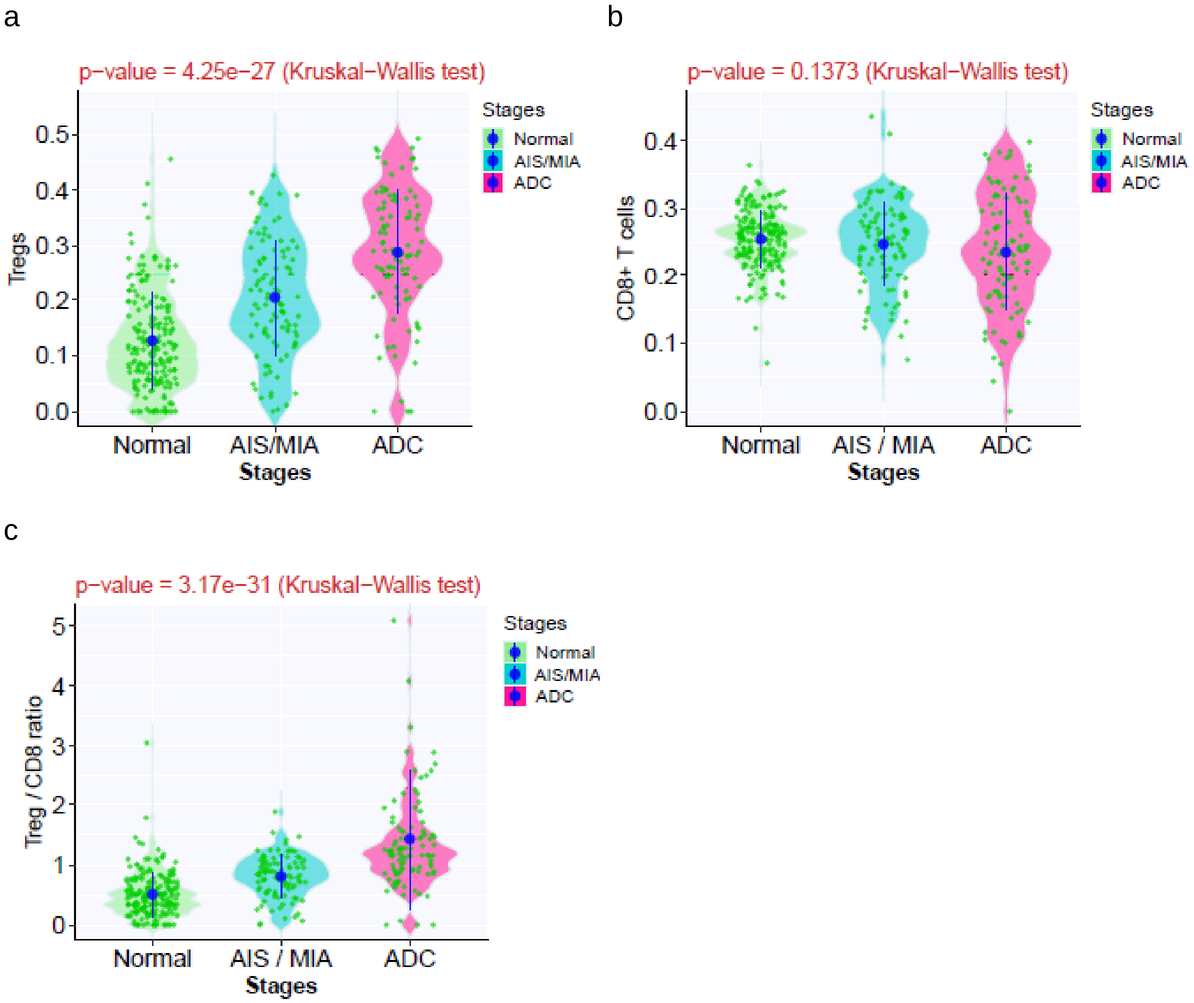


Supplementary Figure 12. Validation for T cell infiltration in IPNs of different stages. The immune cell fraction of T regulatory cells (a), CD8 T-cells (b), Treg/CD8 ratio (c) in normal lung, AIS/MIA and invasive ADC inferred by deconvolution of transcriptomic data from a previously published cohort using ImmuCellAI. Each green dot represents immune cell fraction in each IPN and the blue dots represent the means of immune cell fraction in IPNs of each histologic stage with 95% confidence intervals as error bars. The differences among all stages were assessed using the Kruskal-Wallis H test.

LEGEND FOR SUPPLEMENTARY TABLES

Supplementary Table 1. Patient clinical characteristics

Supplementary Table 2. Summary of clinical characteristics associated with different specimens

Supplementary Table 3. DNA motifs associated with DMRs enriched in lung ADC and its precursors of different stages

Supplementary Table 4. DNA motifs significantly matched to known transcription factor (TF) binding sites

Supplementary Table 5. Transcription factor binding sites and histone marks significantly enriched in chromosomal regions with eloci in each stage

Supplementary Table 6. Clinical, genomic and epigenomic features in IPNs with more than 4 multi-region samples

Supplementary Table 7. Putative tumor suppressor genes showing evidence of convergent genomic and epigenomic evolution
